# Comprehensive analysis of the genetic variation in the *LPA* gene from short-read sequencing

**DOI:** 10.1101/2024.03.21.24304527

**Authors:** Raphael O. Betschart, Georgios Koliopanos, Paras Garg, Linlin Guo, Massimiliano Rossi, Sebastian Schönherr, Stefan Blankenberg, Raphael Twerenbold, Tanja Zeller, Andreas Ziegler

## Abstract

Lipoprotein (a) [LP(a)] is a risk factor for cardiovascular diseases and mainly regulated by the complex LPA gene. We investigated the types of variation in the LPA gene and their predictive performance on LP(a) concentration. We determined the Kringle IV-type 2 (KIV-2) copy number (CN) using the DRAGEN LPA Caller (DLC) and a read-depth based CN estimator in 8351 whole genome sequencing samples from the GENESIS-HD study. The pentanucleotide repeat in the promoter region was genotyped with GangSTR and ExpansionHunter. LP(a) concentration was available in 4861 population-based subjects. Predictive performance on LP(a) concentration was investigated using random forests. The agreement of the KIV-2 CN between the two specialized callers was high (r=0.9966; 95% confidence interval [CI] 0.9965–0.9968). Allele-specific KIV-2 CN could be determined in 47.0% of the subjects using the DLC. Lp(a) concentration can be better predicted from allele-specific KIV-2 CN than total KIV-2 CN. Two single nucleotide variants 4925G>A and rs41272114 further improved prediction. The genetically complex LPA gene can be analyzed with excellent agreement between different callers. The allele-specific KIV-2 CN is more important for predicting LP(a) concentration than the total KIV-2 CN. It would be important that the allele-specific KIV-2 CN is determinable in all subjects.

## Introduction

High LP(a) levels are linked to increased cardiovascular disease (CVD) risk, such as myocardial infarction[1]. LP(a) is a lipoprotein consisting of a large apolipoprotein(a) molecule and a low-density-lipoprotein (LDL) particle [2]. LP(a) shows a broad range of concentrations, ranging from 0.1 mg/dL to more than 200 mg/dL [3,4]. LP(a) levels differ between populations of different geographic origin, and African populations show 2-3-fold higher LP(a) levels than European and Asian populations [4].

LP(a) levels are highly heritable, and the LPA gene explains up to 90% of its variation [3]. The main driver for the variability of LP(a) levels is the Kringle IV (KIV) structure, which consists of 10 subtypes. The subtype KIV-2 is 5.5 kilobases long, highly variable, and the number of KIV-2 repeats ranges from 1 to > 40 [4]. A high number of KIV-2 repeats is associated with low LP(a) protein levels, and more than 95% of sub-jects are heterozygous for the number of KIV-2 repeats [3]. It is important to note that LP(a) protein levels are primarily determined by the number KIV-2 repeats on the shorter allele [5].

The number of KIV-2 repeats has been traditionally measured by laborious wet-lab experiments, such as immunoblotting and pulsed-field gel electrophoresis (PFGE) [6,7]. Because of the strong linkage disequilibrium (LD) between the KIV-2 repeat and several single nucleotide variation (SNV) in the LPA gene, SNVs have been used as simple-to-use proxies for the KIV-2 repeat [4,8,9]. While this approach is accurate for European-like populations, it is diminishing for other populations, especially East Asian-like populations [9].

Recently, a specialized caller for the number of KIV-2 repeats, termed DRAGEN LPA Caller, was released, which allows determining the number of KIV-2 repeats from short-read whole-genome sequencing (WGS) data [10]. This caller estimates the total number of KIV-2 repeats. And depending on the genotypes at two SNVs at position 296 and 1264 within the KIV-2 repeat unit, the caller may be able to determine the allele-specific KIV-2 repeat number [10]. A more general caller, termed read depth-based copy number estimator (CNE), to determine all multicopy genes was publicly released late 2023 [11]. CNE uses the read depth from short-read WGS to estimate the total copy number for the genes of interest.

Several SNVs have been identified which statistically influence LP(a) levels by genome-wide association studies (GWAS) [6,12]. These SNVs are primarily located in the LPA gene. Some SNVs are located in the KIV-2 repeat, and because of its complex structure, a specialized caller is needed. This caller is able to determine if a subject either carries a variant among all KIV-2 repeats or all repeats do not contain the variant [7].

It was suggested that a pentanucleotide repeat (PNR) in the promotor region up-stream of the LPA gene alters LP(a) levels [4,13,14]. Specifically, the PNR alleles with 10 or 11 repeats are associated with a small number of KIV-2 repeats, but counterintuitively express very low LP(a) concentrations [13]. However, it has been demonstrated that the association between a high number of KIV-2 repeats and low LP(a) levels is mediated by a SNV located in the KIV-2 repeat [15]. Therefore, adding this SNV to a model predicting LP(a) concentrations should abolish the association with the PNR.

The aim of the study was to investigate all types of genetic variation in the LPA gene and their association with LP(a) levels. To this end, we used the 8351 WGS from the GENEtic SequencIng Study Hamburg-Davos (GENESIS-HD) study. In 4861 of these subjects LP(a) protein level measurements were available. We compared both callers, DRAGEN LPA Caller and CNE, to determine the number of KIV-2 repeats against each other on their agreement in determining the total number of KIV-2 re-peats. Furthermore, we compared both PNR callers ExpansionHunter and GangSTR and their agreement in determining the number of PNRs. We developed a random forest model to predict LP(a) levels from all types of genetic variation and demonstrate the importance of the number of allele-specific KIV-2 repeats. We also show that the PNR is not predictive for LP(a) measurements, if SNV 4925G>A, located in the KIV-2 repeat, is included.

## Methods

### Cohort

The GENESIS-HD study is a collaborative effort. It was planned to sequence 9000 individuals in total, of which approximately 8000 were to come from the German population-based Hamburg City Health Study (HCHS); for a description of the HCHS design, see [16]. An additional set of approximately 1000 subjects were to be selected from patient-based clinical cohorts with distinct cardiovascular characteristics. The clinical cohorts included subjects with myocardial infarction at young age among others; for details see Betschart et al. (unpublished). In this study, only LP(a) levels measured in 4861 individuals from HCHS were used.

The local ethics committee of the Landesärztekammer Hamburg (State of Hamburg Chamber of Medical Practitioners, PV5131) had no objections against the conduct of the study. The Data Protection Commissioner of the University Medical Center of the University Hamburg-Eppendorf and the Data Protection Commissioner of the Free and Hanseatic City of Hamburg approved the study. The study is registered at ClinicalTrial.gov (NCT03934957).

### Measurement of LP(a)

LP(a) was centrally measured in thawed serum samples previously stored at -80° degrees Celsius with the molar-based Tina-quant® Gen 2 Lipoprotein (a) assay on a COBAS INTEGRA® 400 plus analyzer (Roche Diagnostics, Rotkreuz, Switzerland) at the University Medical Center Hamburg-Eppendorf. Concentrations were reported in nmol/L with a limit of detection (LOD) of 7 nmol/L. The assays inter coefficient of variation (CV) was 5,86%, the inter CV was 3,76%.

### Sequencing

Core elements of the sequencing protocol for the GENESIS-HD study are as follows: After arrival of the DNA samples from the University Medical Center Eppendorf (UKE) in Hamburg at the University Medical Center Zurich (USZ), DNA concentration was measured with PicoGreen, followed by an automated DNA normalization with Hamilton Robotics. The library was constructed according to Illumina TruSeq DNA PCR Free Library Prep protocol HT (Illumina Inc., San Diego, CA, USA) for whole genome sequencing. Briefly, the protocol steps were: 1) fragmentation of 1 μg genomic DNA to 350 bp inserts by Covaris LE220-plus, 2) cleanup of fragmented DNA, 3) re-pair ends, 4) removal of large and small DNA fragments, 5) 3’-end adenylation, and 6) adapter ligation. The resulting library was quantified and quality-assessed with the iSeq100 (Illumina). Samples were normalized according to the quantification values, and 54 samples were pooled for sequencing on an Illumina NovaSeq 6000 sequencer. Samples were sequenced twice on S4 flow cells with 300 cycles (2 × 150 reads) with an estimated coverage of 15 × each, following Illumina protocols. The aimed sequencing depth was that ≥ 95% of all samples had a coverage of ≥ 30 x.

### Pre-processing, quality control and multi-sample calling of WGS data

Pre-processing and quality control (QC) of WGS data has been described else-where Betschart et al. (unpublished). In brief, QC was continuously performed during the conduct of the study, and approximately 600 samples were monthly processed on the used single NovaSeq 6000 sequencer. During normal operation, data were transferred in batches of approximately 250 subjects, and QC reports were generated for these batches. For pre-processing, we used DRAGEN version 3.8.4 on all samples for mapping and alignment and for single sample variant calling, using the hg38 human reference genome. Pre-processing was done without adapter trimming, and read length was 151bp. During QC, PC coefficients were estimated using PCs provided by the 1000Genomes project phase 3 data [17]. The first two estimated PCs were used to define a genetically similar European-like population; for details, see Betschart et al. (unpublished). For the multi-sample calling (also termed joint-calling), the iterative gVCF genotyper has been used.

### Measurement of number of KIV-2 repeats

The total number of KIV-2 repeats was determined with two approaches, the CNE [11] and the DRAGEN LPA Caller [10].

### Read depth-based copy number estimator (CNE)

The CNE software divides the genome into non-overlapping bins with a size of 100bp to estimate GC content and read depth for every sample. Read depth was estimated with mosdepth version 0.3.6 [18]. Because CNE produces relative copy numbers, the output was multiplied by six to account for the number of KIV-2 copies within the hg38.

### DRAGEN LPA Caller

The Illumina DRAGEN LPA Caller is available with DRAGEN version 4.2 [10]. Specifically, we used the DRAGEN version 4.2.0-673-g9e903543. This caller counts reads which fall within the KIV-2 repeat. A total of 3000 additional regions, each with a length of 2000 bases are used for normalization. The read counts for all regions are normalized by their length and by GC-content. The number of KIV-2 repeats is derived from the normalized KIV-2 coverage multiplied by six to account for the number of KIV-2 copies within the reference genome.

To determine the allele-specific number of KIV-2 repeats, the DRAGEN LPA Call-er measures the proportion of reads at two SNVs in linkage disequilibrium (LD) at positions 296 and 1264, which occur in every KIV-2 repeat. The DRAGEN LPA Caller re-ports the number of KIV-2 repeats for both the reference and the alternative allele. The shorter allele is termed KIV-2 short, and the longer allele is named KIV-2 long. We only used the number of KIV-2 repeats determined by the DRAGEN LPA Caller in association analysis because of its ability to determine the number of allele-specific KIV-2 re-peats.

### Analysis of the pentanucleotide repeat (PNR)

The number of repeats of the PNR were determined with GangSTR version 2.5.0 [19] and ExpansionHunter version 5.0.0 [20], both with default parameters. The ge-nomic location of the PNR is chr6:160665585-160665629 in hg38. Oketch et al. [21] demonstrated good agreement between these two short tandem repeat callers and showed fewer Mendelian inheritance errors of GangSTR in the analysis of Genome in a Bottle trios [22]. The resulting variant calling files (VCFs) were parsed for the REPCN field with bcftools query version 1.18 [23]. If a subject was heterozygous for the number of PNRs, the lower number (shorter allele) was assigned to the variable Allele 1, and the higher number was assigned to Allele 2 [15]. The polyserial correlation be-tween the number of PNR and the carrier status of KIV-2 SNV 4925G>A was estimated using the polyserial function from the polycor R package.

### Analysis of the KIV-2 single nucleotide variations (SNVs)

SNVs within the KIV-2 repeat were genotyped with the pipeline termed exome vntr nf [7]. The pipeline takes aligned reads as input (BAM file) and extract the reads that fall within the LPA genomic region, i.e., chr6:160530484-160665259 in hg38. These reads are then converted to a FASTQ file and realigned to a single KIV-2 repeat. In the last step, the variants are called with mutserve, a specialized caller originally developed for mitochondrial variant calling [24]. This caller only provides information on carrier status, which is defined as either carrier or non-carrier.

### Statistical analysis

#### Descriptive statistics

Descriptive characteristics of important variable are provided with median and quartiles. For dichotomous variables, absolute and relative frequencies are given. Agreement between the two KIV-2 callers was determined using Pearson correlation, a scatterplot, and a Bland-Altman plot. Agreement between the two PNR callers was determined using Pearson correlation and dotplot.

#### Genome-wide association study (GWAS)

GWAS for the LP(a) concentration was performed with REGENIE version 3.2.5.3 [25]. A minor allele frequency (MAF) threshold of 0.005 and a Hardy-Weinberg equilibrium (HWE) threshold of 10-9 were used for filtering diallelic SNVs. Subjects showing a kinship coefficient of 3rd cousins or closer (≥ 0.044; [26]) were partitioned to create an unrelated subset with the highest number of unrelated subjects using the pcairPartition function from the GENESIS R package version 2.28.0 [27].

For association analyses, LP(a) measurements were log-transformed. An additive genetic model was used, and adjustments were done for age, sex and the first five principal components (PCs). The genome-wide significance threshold was set to 5 × 10-8. Genotypes of all statistically significant SNVs without missing values were extracted with the R package SeqVarTools version 1.40.0 [28].

#### Predictive model for LP(a) levels using random forests

Because of the high LD between the different genetic markers in the LPA gene, i.e., high multicollinearity, we used random forests by employing the ranger R package [29] to investigate the importance of multiple genetic markers on LP(a) levels.

In the first step, we estimated two random forests. Random forest 1 (RF1) included all genetic variation plus the allele-specific number KIV-2 repeats. Random forest 2 (RF2) included all genetic variation plus the total number of KIV-2 repeats. The genetic variation consisted in all SNVs from the GWAS, all SNVs from the specialized SNVs caller in the KIV-2 repeat and the number of PNRs estimated by GangSTR. To estimate the variables with large contribution to the predictive performance, we estimated the conditional predictive impact (CPI) using the CPI R package. We used default values for the CPI function [30]. The CPI tests for conditional independence and measures the variable importance. Specifically, the CPI of a variable provides the information how much the predictive performance deteriorates if the variable was replaced by a non-informative variable. CPIs, their CIs and test statistics were estimated from 10-fold cross-validation (CV). Variables with a CPI p-value < 0.05 were kept.

For all random forests, we tuned the following hyperparameters using 10-fold nested CV: minimal node size (min.node.size), percentage of included variables in each splitting step (mtry.ratio), and tree depth (max.depth). Default values were used for other parameters. Hyperparameter tuning was done with the mlr3 R package (version 0.17.2) [31]. The hyperparameters of the best performing model, i.e., the one with the lowest root mean square error (RMSE), was then used in the CPI calculations.

The direct comparison of the predictive performance between the number of allele-specific KIV-2 repeats and the total number KIV-2 repeats was done with the following model:

1. Full model: inclusion of all available genetic variation plus
  a. the total number of KIV-2 repeats
  b. the allele-specific number of KIV-2 repeats
2. KIV-2 RF1: inclusion of genetic variation with CPI p < 0.05 from RF1 plus
  a. the total number of KIV-2 repeats
  b. the allele-specific number of KIV-2 repeats
3. KIV-2 RF2: inclusion of genetic variation with CPI p < 0.05 from RF2 plus
  a. the total number of KIV-2 repeats
  b. the allele-specific number of KIV-2 repeats

Since the random forests for the total number of KIV-2 repeats and the allele-specific number of KIV-2 repeats are estimated from the same CV data, they are directly comparable. Our hypotheses were:

- Hypothesis 1: The full model shows the highest predictive performance
- Hypothesis 2: RF1 is sparser than RF2
- Hypothesis 3: Model KIV-2 RF1 b performs better than KIV-2 RF1 a
- Hypothesis 4: Model KIV-2 RF2 b performs better than KIV-2 RF2 a
- Hypothesis 5: Full models a and b show similar performance, because proxy SNVs from the LPA gene should compensate for the additional information for the allele-specific number of KIV-2 repeats

The global significance level was set to 0.05. To adjust for multiple testing, hypotheses were tested hierarchically, starting with hypothesis 1. For hypothesis 2, no formal statistical test was performed. For all models, we forced sex and age to be available for splitting in all trees and all splitting steps using the ranger option always.split.variable [29]. If the final model does not contain age or sex, the coefficient of determination (R^2^) obtained from this model can be interpreted as locus-specific heritability. R^2^ was estimated for all models.

#### Software and hardware

The statistical analysis was performed with R version 4.3.2 [32] on our on-premises HPC equipped with 4 computing nodes. Each node is equipped with 2 AMD EPYC 7742 CPUs and a total of 2TB of RAM. The entire R workflow was run with targets version 1.3.2 [33]. Plots were generated with ggplot2 version 3.4.4 [34]. The code for the analyses is provided as a supplement.

## Results

### Study characteristics

Characteristics of GENESIS-HD subjects with available LP(a) measurements are displayed in Table 1. Figure 1 shows the LP(a) concentration per binned total number of KIV-2 repeats of GENESIS-HD subjects with available LP(a) measurements. LP(a) concentration per binned number KIV-2 repeats on the short and long allele are provided in Figures S1 and S2, respectively.

**Table 1.** Characteristics of GENESIS-HD subjects with available LP(a) measurements. Continuous characteristics are provided as median and interquartile range (IQR), dichotomous variables as number of subjects and percentage.

| Demographics | N = 4861 |
| --- | --- |
| Age [years] | 63 (56, 70) |
| Sex [male] | 2425 (49.9%) |
| LP(a) [nmol/L] | 16.7 (7.50, 59.0) |
| Total number of KIV-2 repeats | 39.0 (34.1, 43.9) |
| Number of KIV-2 repeats long allele | 22.7 (20.0, 25.0) |
| Number of KIV-2 repeats short allele | 16.6 (13.4, 19.9) |
| Allele-specific number of KIV-2 repeats available | 2274 (47.0%) |
| 4925G>A carrier | 1124 (23.1%) |
| rs41272114 carrier | 283 (5.8%) |

**Figure 1.**
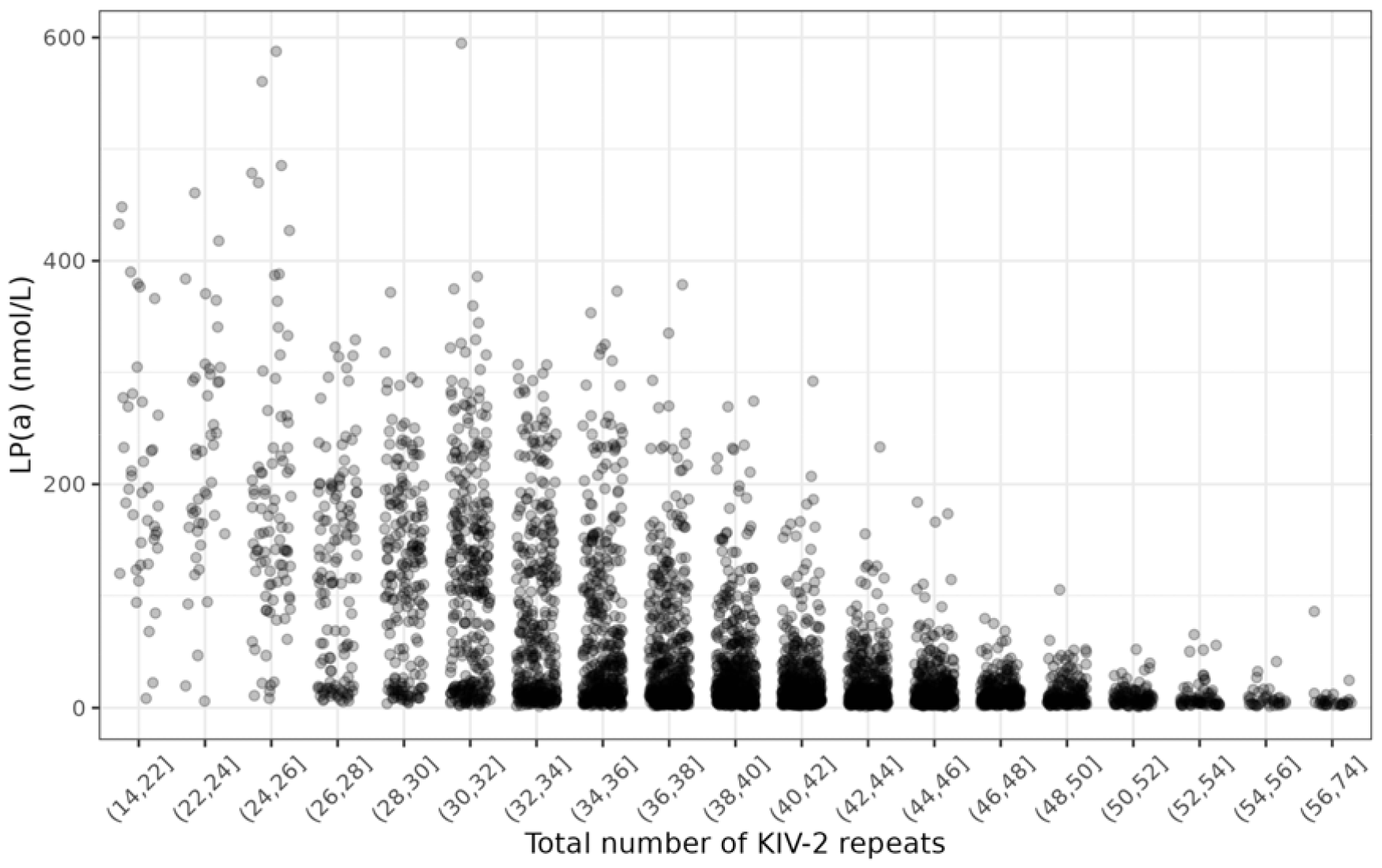
Scatterplot for LP(a) and total number of KIV-2 repeats (n = 4861). KIV-2 CNs were binned so that there are at least 30 samples in each bin. Horizontal jittering was added to prevent overlapping between points.

### Agreement between specialized variant callers

Figure 2 displays the scatterplot for the total number of KIV-2 repeats estimated by the DRAGEN LPA Caller and the CNE. The correlation was 0.9966 (CI 0.9965-0.9968). Figure S3 shows the corresponding Bland-Altman plot.

**Figure 2.**
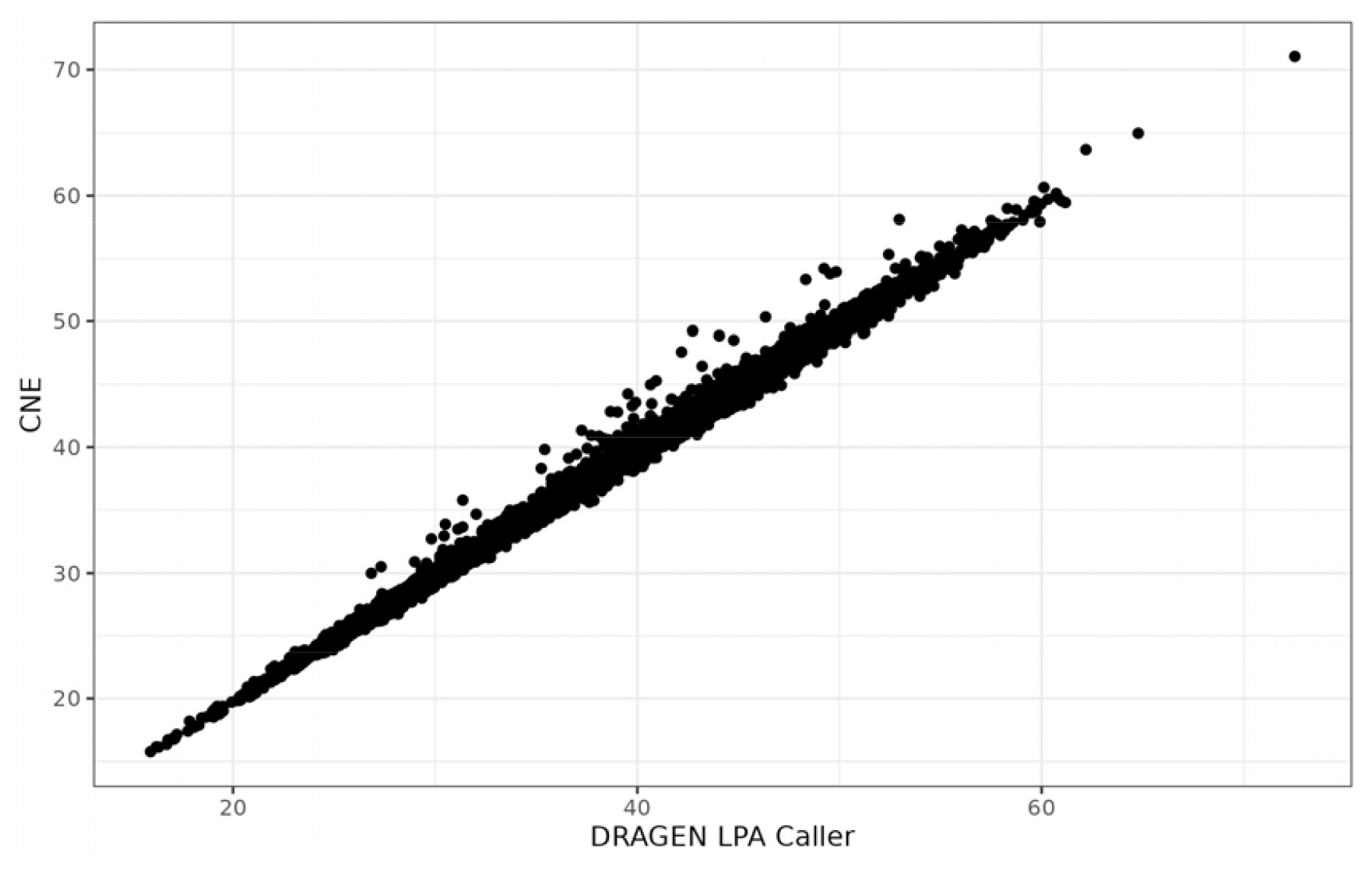
Scatterplot for total number of KIV-2 repeats estimated by the DRAGEN LPA Caller and read depth based CN estimator (CNE) (n = 8351).

Both GangSTR and ExpansionHunter successfully estimated the genotypes of the PNR in the promoter region of the *LPA* gene on all 8351 subjects. The Pearson correlation between the callers was 0.9938 (CI 0.9936-0.9941) for allele 1 and slightly higher for allele 2 (r = 0.9961, CI 0.9960-0.9963). Figure S4 shows the dotplot for the number of subjects per genotype. In agreement with [1], we observed a high LD between the number of PNR and 4925G>A with a polyserial correlation of 0.991.

### Agreement of allele-specific number of KIV-2 repeats with results from 1000 Genomes Project

We compared the number of KIV-2 repeats of the GENESIS-HD dataset with the 1000 Genomes dataset (1KGP) analysed by Behera et al. [2]. In 3925 out of the 8351 subjects (47.0%), the DRAGEN LPA Caller determined the allele-specific number of KIV-2 repeats. Excellent agreement between the GENESIS-HD dataset and the European ancestral population (633 samples) could be observed (Figure 3)

**Figure 3.**
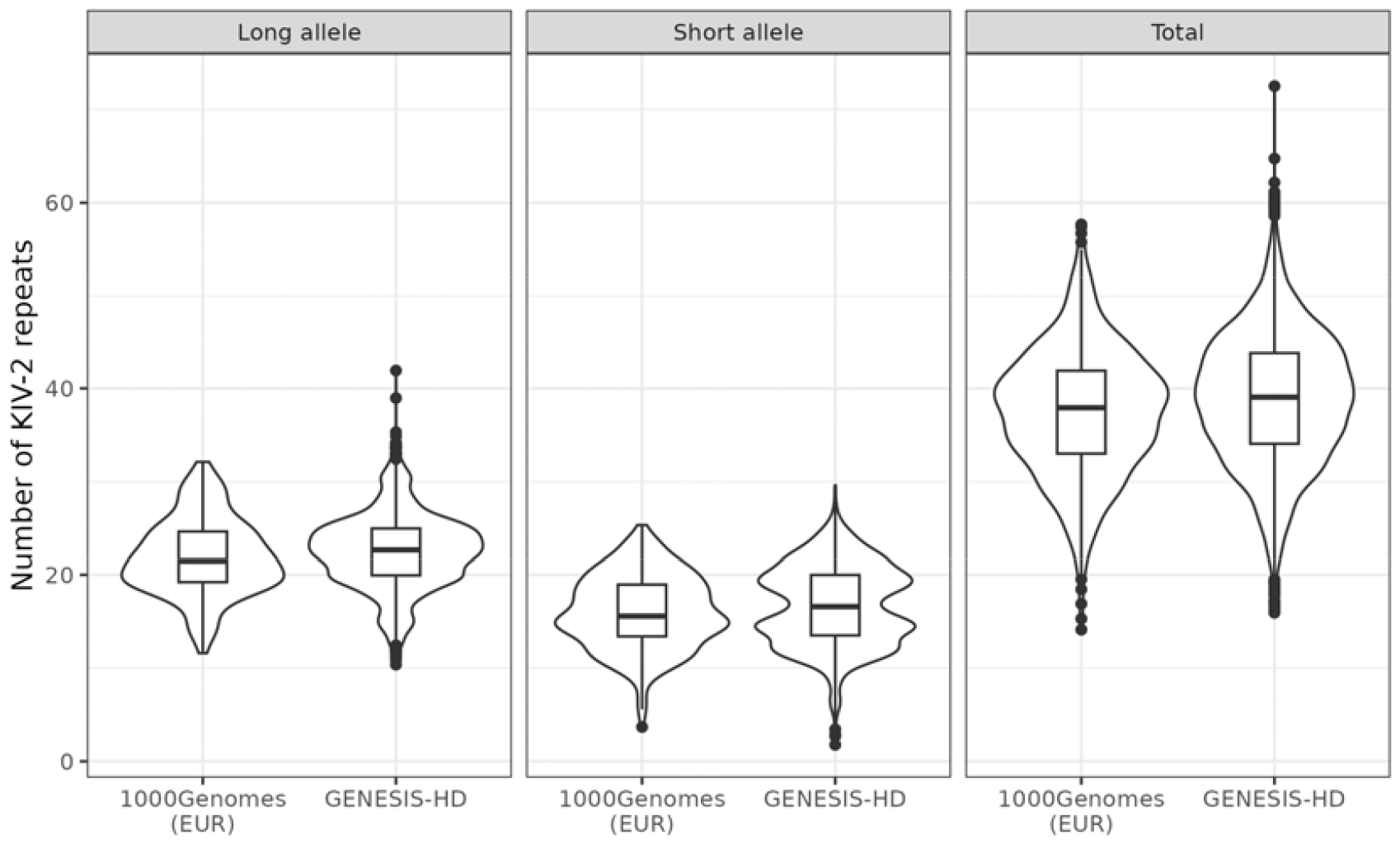
Violin and boxplots of number of KIV-2 repeats for GENESIS-HD samples and EUR-like samples (EUR) from 1000 Genomes project phase 3 (1KGP) [3] (n = 633). For the total number of KIV-2 repeats, 8351 subjects were included. For the number of KIV-2 repeats on the short and long allele, 3925 subjects were included.

### Genome-wide association study for LP(a) concentration

Filtering and quality control (QC) left us with a total of 10.3 million (10,334,912) diallelic SNVs in 4803 unrelated subjects. The only genome-wide significant region for LP(a) concentration was the *LPA* gene (Figure S5). A total of 855 SNVs were found to be statistically significant in the *LPA* gene (hg38: chr6:160531482-160664275) with a padding of 500kb (Table S1). The lead SNV was rs56393506 (MAF: 0.16; per-allele change in LP(a) concentration: 1.065 [95% CI 0.997-1.132]; p=1.74 × 10^−211^; for locus zoom plot, see Figure S6). The scaled genomic inflation factor was 0.995.

### Genetic variants selected by conditional predictive impact (CPI)

Four and six variables were selected by conditional predictive impact for the allele-specific KIV-2 random forest model RF1 and the total KIV-2 random forest model RF2, respectively (Table 2). In both models, the number of KIV-2 repeats had the highest conditional predictive impact. In the allele-specific KIV-2 model, the shorter allele had a stronger effect on LP(a) levels than the longer allele. Two SNVs 4925G>A and rs41272114 had a significant conditional predictive impact in both models, and the contingency table is provided in Table 3.

**Table 2.** Variables selected by conditional predictive impact (CPI) for both random forest models RF1 and RF2. CPIs, their 95% confidence intervals (95% CI) and corresponding p-values are displayed. CPIs and 95% CIs were multiplied by 10^3^.

| Variable | RF1 (allele-specific) |  |  | RF2 (total) |  |  |
| --- | --- | --- | --- | --- | --- | --- |
|  | CPI | 95% CI | p-value | CPI | 95% CI | p-value |
| Short KIV-2 | 97.7 | 77.8–117.7 | $5.5 \times 10^{-16}$ | – | – | – |
| Long KIV-2 | 22.6 | 14.3–30.9 | $3.8 \times 10^{-6}$ | – | – | – |
| Total KIV-2 | – | – | – | 58.9 | 43.6–70.1 | $1.2 \times 10^{-10}$ |
| 4925G>A | 9.9 | 4.3–15.5 | $1.8 \times 10^{-3}$ | 8.7 | 2.0–15.3 | $1.6 \times 10^{-2}$ |
| rs41272114 | 3.6 | 1.0–6.3 | $1.3 \times 10^{-2}$ | 4.4 | 0.6–8.1 | $2.7 \times 10^{-2}$ |
| rs9347465 | – | – | – | $9.6 \times 10^{-4}$ | $1.2 \times 10^{-6}$–$1.9 \times 10^{-3}$ | $4.9 \times 10^{-2}$ |
| rs2489959 | – | – | – | $8.6 \times 10^{-4}$ | $5.6 \times 10^{-6}$–$1.7 \times 10^{-3}$ | $4.9 \times 10^{-2}$ |
| rs2457567 | – | – | – | $8.4 \times 10^{-4}$ | $3.2 \times 10^{-5}$–$1.6 \times 10^{-3}$ | $4.4 \times 10^{-2}$ |

**Table 3.**
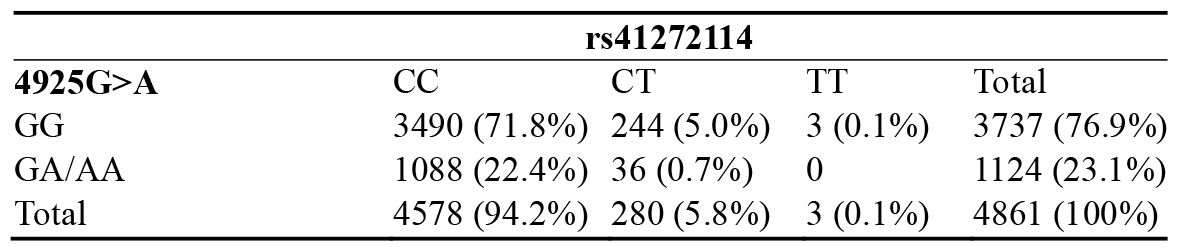
Contingency table for SNV 4925G>A carrier status and rs41272114.

Non-carries of both SNVs (4925GG and rs41272114CC; n=3488), had a median LP(a) concentration of 20.3 nmol/L (7.3, 83.0). Subjects carrying at least one A allele at SNV 4925 and genotype CC at rs41272114 (n=1088) showed a lower LP(a) concentration of 15.0 nmol/L (9.7-29.3). The lowest median LP(a) concentration of 4.8 nmol/L (2.9-15.8) was observed in subjects carrying variant alleles at rs41272114, but not carrying an A allele at SNV 4925 (n=247). Individuals carrying variations at both SNVs (n=36) had an LP(a) concentration of 8.4 nmol/L (6.3-12.7). It should be noted that the number of double carriers is low. The LP(a) concentration of the genotype combination and short and total number of KIV-2 repeats is displayed in Figure 4.

**Figure 4.**
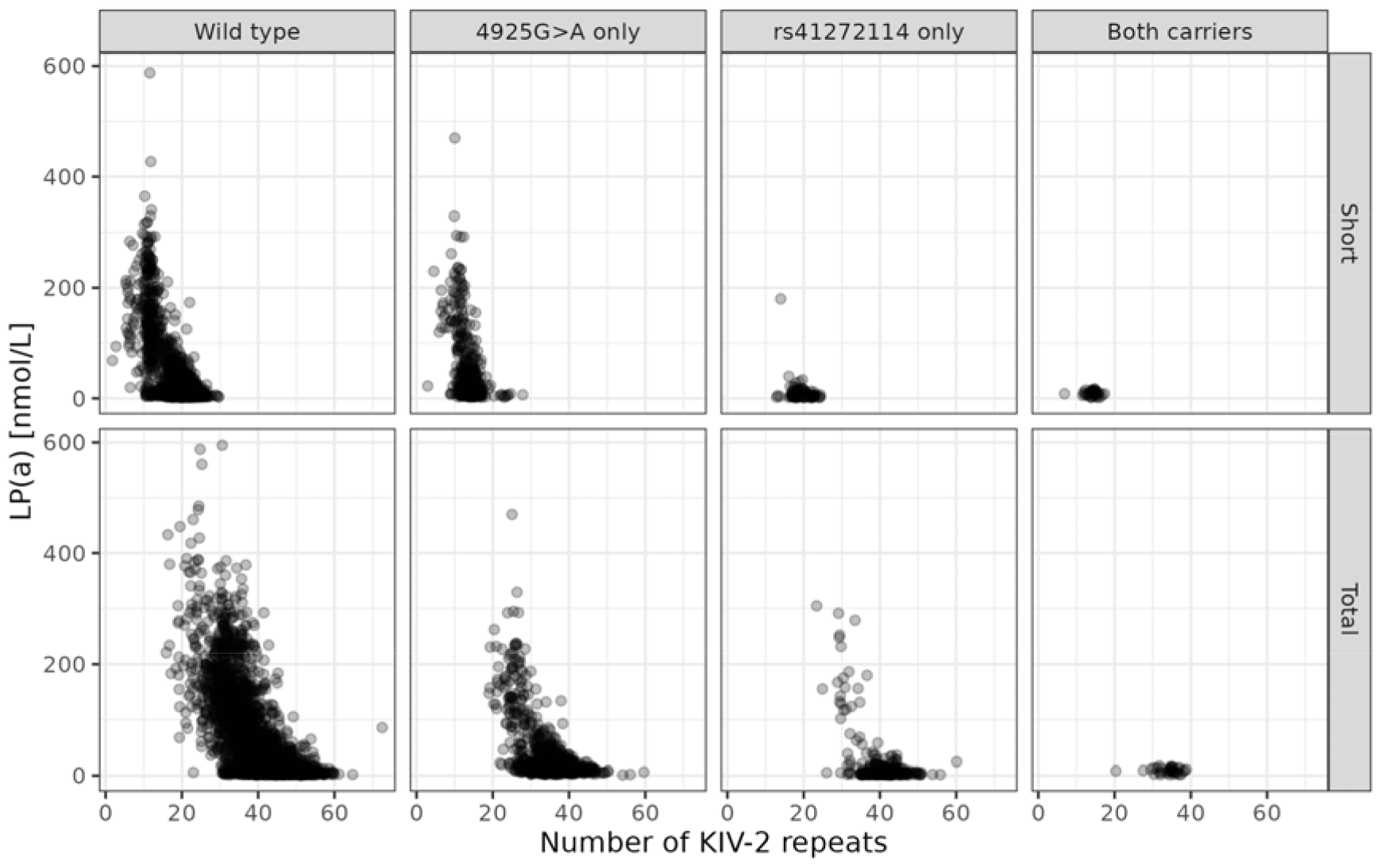
LP(a) concentration by number of KIV-2 repeats for all single nucleotide variation combinations of 4925G>A and rs41272114 carriers of the T allele. Upper panels: number of KIV-2 repeats of the short allele. Lower panels: total number of KIV-2 repeats.

Only three subjects were homozygous TT at rs41272114; these tree subjects did not harbour an A allele at SNV 4925. These subjects had LP(a) concentrations of 2.2, 2.3, and 3.0 nmol/L.

The three SNVs from model RF2 rs9347465, rs2489959, and rs2457567 were neither found in the GWAS catalogue nor by a Medline search. As expected, the PNR did not have a significantly conditional predictive impact.

### Comparison of predictive performances for LP(a) concentrations

Mean and standard deviation (SD) of the difference in R^2^ between allele-specific number and total number of KIV-2 repeats are displayed in Table 4. Specifically, we estimated the difference in R^2^ between the allele-specific number and total number of KIV-2 repeats. The full model showed the highest predictive performance for both the total number of KIV-2 repeats (R^2^=0.6855, CI 0.6109-0.7601) and the allele-specific number of KIV-2 repeats for the total number of KIV-2 repeats (R^2^=0.6953, CI 0.6414-0.7492). The performance of the full model was significantly better than that of RF1 and RF2 (both p < 0.001; Table S2). This confirms hypothesis 1. For model RF1, four variables were selected, compared to six variables for model RF2. RF1 was thus sparser than RF2, which confirms hypothesis 2.

**Table 4.**
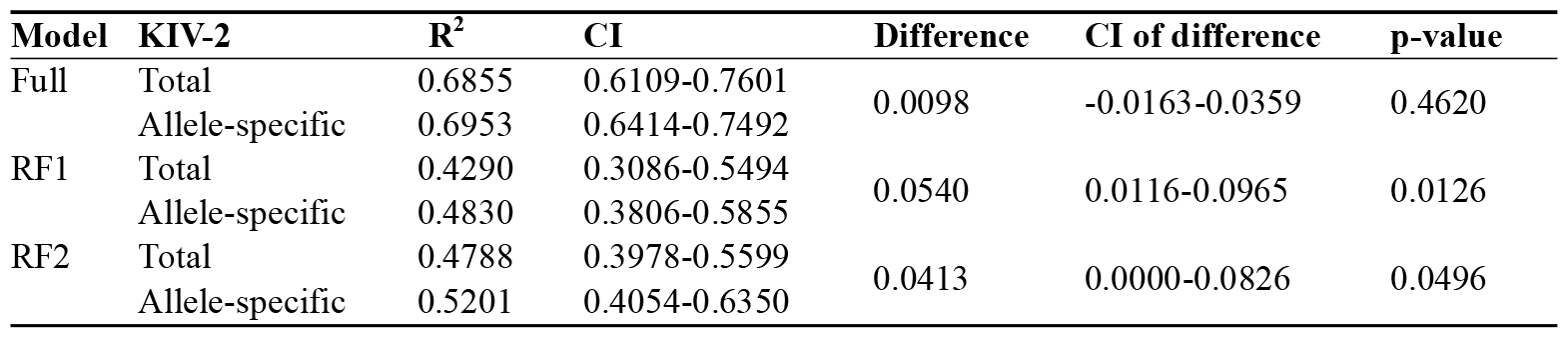
Coefficient of determination (R^2^) and 95% confidence interval (CI) of different random forest (RF) models for LP(a) concentrations.

Model RF1 b showed a significantly better predictive performance than model RF1 a (p=0.0126), which confirms hypothesis 3. Similarly, model RF2 b outperformed model RF2 a (p=0.0496), which confirms hypothesis 4. Finally, no statistically significant difference was observed between the total and allele-specific KIV-2 repeat models in the full model (p=0.4620), which is in agreement with hypothesis 5.

Full model: All available genetic variation; RF1: inclusion of genetic variation with conditional predictive impact (CPI) p < 0.05 from allele-specific model, when allele-specific numbers of KIV-2 repeats were available; RF2: inclusion of genetic variation with CPI p < 0.05 from total model, when total number of KIV-2 repeats was available.

## Discussion

In this study, we performed a comprehensive analysis of the genetic variation within the *LPA* gene by using data from whole genome short-read sequencing. Key to these analyses was the availability of specialized callers to estimate the number of KIV-2 repeats. There was excellent agreement between the DRAGEN LPA Caller and the CNE, which were both publicly released in 2023. The accuracy of the DRAGEN LPA Caller was previously confirmed by long-read sequencing [2], and the present analysis indirectly confirms the validity of the CNE caller for the *LPA* gene.

A weakness of both callers is the estimation of the number of allele-specific KIV-2 repeats. This information is important, because the number of allele-specific KIV-2 repeats leads to better predictive performance for LP(a) concentration. Of note, the CNE is able to estimate the total number of KIV-2 repeats only. In contrast, the DRAGEN LPA Caller is able to estimate the number of allele-specific KIV-2 repeats, depending on the genotypes of two SNVs within the KIV-2 region. In this study, the DRAGEN LPA caller provided the number of allele-specific KIV-2 repeats in 47% of all subjects, which were of European ancestry. This percentage varies between subjects of different ancestry groups. For example, in AMR-like subjects, the percentage was the lowest with 38%, and highest in AFR-like individuals with 52% [2].

There was a high agreement between the two callers ExpansionHunter and GangSTR for estimating the number of PNRs. However, the genetic information of the PNR is not important for predicting LP(a) concentrations because almost all of the genetic information of the multiallelic PNR is captured by the dichotomous SNV 4925G>A, located within the KIV-2 repeat (correlation = 0.997). This is in agreement with previously published results [1].

The results of our GWAS confirmed previous reports that variants associated with LP(a) concentrations are primarily located in the *LPA* gene region [4]. We identified 855 significant SNVs surrounding the *LPA* gene. The lead SNV rs56393506 has been previously reported as being associated with increased LP(a) concentration [5].

Predicting LP(a) concentrations based on genetic variation is challenging because of high LD in the *LPA* gene. This high multicollinearity between genetic variation invalidates classical regression models, such as linear regression or quantile regression. In contrast, some of the machine learning methods, such as random forests, are able to adequately deal with a high LD. The random forest models revealed that only a small number of genetic variation had a significant impact on the predictions of the LP(a) concentrations. The full model incorporating all genetic variation from the *LPA* gene had a substantially higher predictive performance of LP(a) concentrations, when compared to models which covered the most important genetic markers (RF1 and RF2). Indeed, the locus-specific heritability (R) of the full model was 0.83 (CI 0.82– 0.84), while it was only 0.68 (CI 0.65–0.70) and 0.71 (CI 0.69–0.72) for RF1 and RF2, respectively. The locus-specific heritability of the full model was thus in the same magnitude as that of other studies [6, 7].

In line with the literature [6, 7], the number of KIV-2 repeats had the highest conditional predictive impact, with the number of allele-specific KIV-2 repeats being more predictive than the total number of KIV-2 repeats. The two most important SNVs for predicting LP(a) concentrations were 4925G>A and rs41272114, which confirms other work [6]. SNV 4925G>A is located on a splice site within the KIV-2 repeat and leads to a decrease in LP(a) concentration [7-9]. rs41272114 is also located on a splice site, and T alleles at this SNV result in null alleles [4, 6, 8]. Such null alleles lead to extremely low LP(a) concentrations, which we also observed in this study. Three subjects were homozygous TT at rs41272114. The highest LP(a) concentration is these individuals was 3.0 nmol/L, which is in the bottom five percent of the LP(a) concentration levels among all subjects.

A limitation of our study is that analysis were based on a specific assay to measure the LP(a) concentration. Specifically, we used the Roche assay, which reports the molar LP(a) concentration (nmol/L). Various studies have reported differences between LP(a) assays [10, 11]. Furthermore, we restricted our study to EUR-like subjects only. Future research should investigate the predictive performance of all genetic variation within the LPA gene in different ancestry groups.

The use of long-read sequencing would have enabled us to not only determine the allele-specific number of KIV-2 repeats, but also the genetic variation within the KIV-2 region. However, even long-read technologies show non-unique alignments in the KIV-2 region [2]. The longer the reads, the lower the fraction of non-unique alignments. Furthermore, time and cost for such an endeavour in thousands of subjects are currently beyond reach.

In the future, it would be of great importance to determine the number of allele-specific KIV-2 repeats in all subjects, across all different genetic ancestry groups. Furthermore, determining not only the carrier-status of SNVs within the KIV-2 repeats but also the genotype could lead to better understanding of the complex *LPA* gene.

## Author Contributions

Conceptualization, R.O.B. and A.Z.; data curation, R.O.B., L.G., R.T. and T.Z.; formal analysis, R.O.B; funding acquisition, S.B., R.T., T.Z. and A.Z.; investigation, S.B., T.Z., R.T., and A.Z.; methodology, R.O.B. and A.Z.; project administration, A.Z.; resources, S.B., R.T., T.Z. and A.Z.; software, R.O.B, G.K., P.G., M.R. and S.S.; supervision, A.Z.; visualization R.O.B. and G.K.; writing—original draft, R.O.B. and A.Z; writing—review & editing, All. All authors have read and agreed to the published version of the manuscript.

## Funding

We gratefully acknowledge funding of the GENESIS-HD study by the Kühne Foundation and the measurement of Lp(a) in HCHS by Amgen.

## Data Availability Statement

The data used in this study may not be shared due to privacy issues. The targets file used for the analysis is available in the supplement.

## Acknowledgments

We are grateful to Patricia Bartoschek, Satya Bhowmik, Anna Lena Engels, Tim Hartmann, Yumi Hartmann, Lilia Kisselmann, Anna-Lena Post, and René Riedl for excellent laboratory work with DNA extraction, data management, and quality control.

## Conflicts of Interest

M.R. is an employee of Illumina. R.T. holds a professorship in clinical cardiology at the University Medical Center Hamburg-Eppendorf, supported by the Kühne Foundation and reports research support from the German Center for Cardiovascular Research (DZHK), the Joachim Herz Foundation, the Swiss National Science Foundation (Grant No P300PB_167803), and the Swiss Heart Foundation as well as speaker honoraria/consulting honoraria from Abbott, Amgen, Astra Zeneca, Psyros, Roche, Siemens, Singulex and Thermo Scientific BRAHMS. T.Z. is supported by the German Research Foundation, the EU Horizon 2020 programme, the EU ERANet and ERAPreMed Programmes, the German Centre for Cardiovascular Research (DZHK, 81Z0710102), and the German Ministry of Education and Research. S.B., R.T., T.Z., and A.Z. are listed as co-inventors of an international patent on the use of a computing device to estimate the probability of myocardial infarction (International Publication NumberWO2022043229A1). R.T. and T.Z. are shareholders of the ART-EMIS Hamburg GmbH. A.Z. is scientific director and R.O.B. and G.K. are employees of Cardio-CARE, which is shareholder of the ART-EMIS Hamburg GmbH.

## Notes

### Funding Statement

We gratefully acknowledge funding of the GENESIS-HD study by the Kuehne Foundation and the measurement of Lp(a) in HCHS by Amgen.

### Author Declarations

The local ethics committee of the Landesaerztekammer Hamburg (State of Hamburg Chamber of Medical Practitioners, PV5131) had no objections against the conduct of the study. The Data Protection Commissioner of the University Medical Center of the University Hamburg-Eppendorf and the Data Protection Commissioner of the Free and Hanseatic City of Hamburg approved the study. The study is registered at ClinicalTrial.gov (NCT03934957).

## References

1. Kamstrup, P.R.; Tybjaerg-Hansen, A.; Steffensen, R.; Nordestgaard, B.G. Genetically elevated lipoprotein(a) and increased risk of myocardial infarction. JAMA 2009, 301, 2331–2339, doi:10.1001/jama.2009.801.

2. Berg, K. A new serum type system in man – the Ld system. Vox Sang 1965, 10, 513–527, doi:10.1111/j.1423-0410.1965.tb01404.x.

3. Kronenberg, F.; Utermann, G. Lipoprotein(a): resurrected by genetics. J Intern Med 2013, 273, 6–30, doi:10.1111/j.1365-2796.2012.02592.x.

4. Schmidt, K.; Noureen, A.; Kronenberg, F.; Utermann, G. Structure, function, and genetics of lipoprotein (a). J Lipid Res 2016, 57, 1339–1359, doi:10.1194/jlr.R067314.

5. Gaubatz, J.W.; Ghanem, K.I.; Guevara, J.; Nava, M.L.; Patsch, W.; Morrisett, J.D. Polymorphic forms of human apolipoprotein[a]: inheritance and relationship of their molecular weights to plasma levels of lipoprotein[a]. J Lipid Res 1990, 31, 603–613, doi:10.1016/s0022-2275(20)42828-7.

6. Mack, S.; Coassin, S.; Rueedi, R.; Yousri, N.A.; Seppala, I.; Gieger, C.; Schonherr, S.; Forer, L.; Erhart, G.; Marques-Vidal, P.; et al. A genome-wide association meta-analysis on lipoprotein (a) concentrations adjusted for apolipoprotein (a) isoforms. J Lipid Res 2017, 58, 1834–1844, doi:10.1194/jlr.M076232.

7. Coassin, S.; Erhart, G.; Weissensteiner, H.; Eca Guimaraes de Araujo, M.; Lamina, C.; Schonherr, S.; Forer, L.; Haun, M.; Losso, J.L.; Kottgen, A.; et al. A novel but frequent variant in LPA KIV-2 is associated with a pronounced Lp(a) and cardiovascular risk reduction. Eur Heart J 2017, 38, 1823–1831, doi:10.1093/eurheartj/ehx174.

8. Coassin, S.; Kronenberg, F. Lipoprotein(a) beyond the kringle IV repeat polymorphism: the complexity of genetic variation in the LPA gene. Atherosclerosis 2022, 349, 17–35, doi:10.1016/j.atherosclerosis.2022.04.003.

9. Mukamel, R.E.; Handsaker, R.E.; Sherman, M.A.; Barton, A.R.; Zheng, Y.; McCarroll, S.A.; Loh, P.R. Proteincoding repeat polymorphisms strongly shape diverse human phenotypes. Science 2021, 373, 1499–1505, doi:10.1126/science.abg8289.

10. Behera, S.; Belyeu, J.R.; Chen, X.; Paulin, L.F.; Nguyen, N.Q.H.; Newman, E.; Mahmoud, M.; Menon, V.K.; Qi, Q.; Joshi, P.; et al. Identification of allele-specific KIV-2 repeats and impact on Lp(a) measurements for cardiovascular disease risk. bioRxiv 2023, doi:10.1101/2023.04.24.538128.

11. Garg, P.; Jadhav, B.; Lee, W.; Rodriguez, O.L.; Martin-Trujillo, A.; Sharp, A.J. A phenome-wide association study identifies effects of copy-number variation of VNTRs and multicopy genes on multiple human traits. Am J Hum Genet 2022, 109, 1065–1076, doi:10.1016/j.ajhg.2022.04.016.

12. Lu, W.; Cheng, Y.C.; Chen, K.; Wang, H.; Gerhard, G.S.; Still, C.D.; Chu, X.; Yang, R.; Parihar, A.; O’Connell, J.R.; et al. Evidence for several independent genetic variants affecting lipoprotein (a) cholesterol levels. Hum Mol Genet 2015, 24, 2390–2400, doi:10.1093/hmg/ddu731.

13. Mooser, V.; P. Mancini, F.; Bopp, S.; Petho-Schramm, A.; Guerra, R.; Boerwinkle, E.; Muller, H.-J.; H. Hobbs, H. Sequence polymorphisms in the apo(a) gene associated with specific levels of Lp(a) in plasma. Hum Mol Genet 1995, 4, 173–181.

14. Prins, J.; Leus, F.R.; Bouma, B.N.; van Rijn, H.J. The identification of polymorphisms in the coding region of the apolipoprotein (a) gene–association with earlier identified polymorphic sites and influence on the lipoprotein (a) concentration. Thromb Haemost 1999, 82, 1709–1717.

15. Grüneis, R.; Weissensteiner, H.; Lamina, C.; Schönherr, S.; Forer, L.; Di Maio, S.; Streiter, G.; Peters, A.; Gieger, C.; Kronenberg, F.; et al. The kringle IV type 2 domain variant 4925G>A causes the elusive association signal of the LPA pentanucleotide repeat. J Lipid Res 2022, 63, 100306, doi:10.1016/j.jlr.2022.100306.

16. Jagodzinski, A.; Johansen, C.; Koch-Gromus, U.; Aarabi, G.; Adam, G.; Anders, S.; Augustin, M.; der Kellen, R.B.; Beikler, T.; Behrendt, C.A.; et al. Rationale and design of the Hamburg City Health Study. Eur J Epidemiol 2020, 35, 169–181, doi:10.1007/s10654-019-00577-4.

17. The 1000 Genomes Project Consortium. A global reference for human genetic variation. Nature 2015, 526, 68–74, doi:10.1038/nature15393.

18. Pedersen, B.S.; Quinlan, A.R. Mosdepth: quick coverage calculation for genomes and exomes. Bioinformatics 2018, 34, 867–868, doi:10.1093/bioinformatics/btx699.

19. Mousavi, N.; Shleizer-Burko, S.; Yanicky, R.; Gymrek, M. Profiling the genome-wide landscape of tandem repeat expansions. Nucleic Acids Res 2019, 47, e90, doi:10.1093/nar/gkz501.

20. Dolzhenko, E.; van Vugt, J.; Shaw, R.J.; Bekritsky, M.A.; van Blitterswijk, M.; Narzisi, G.; Ajay, S.S.; Rajan, V.; Lajoie, B.R.; Johnson, N.H.; et al. Detection of long repeat expansions from PCR-free whole-genome sequence data. Genome Res 2017, 27, 1895–1903, doi:10.1101/gr.225672.117.

21. Oketch, J.W.; Wain, L.V.; Hollox, E.J. A comparison of software for analysis of rare and common short tandem repeat (STR) variation using human genome sequences from clinical and population-based samples. bioRxiv 2023, 2022.2005.2025.493473v493473, doi:10.1101/2022.05.25.493473.

22. Zook, J.M.; McDaniel, J.; Olson, N.D.; Wagner, J.; Parikh, H.; Heaton, H.; Irvine, S.A.; Trigg, L.; Truty, R.; McLean, C.Y.; et al. An open resource for accurately benchmarking small variant and reference calls. Nat Biotechnol 2019, 37, 561–566, doi:10.1038/s41587-019-0074-6.

23. Danecek, P.; Bonfield, J.K.; Liddle, J.; Marshall, J.; Ohan, V.; Pollard, M.O.; Whitwham, A.; Keane, T.; McCarthy, S.A.; Davies, R.M.; et al. Twelve years of SAMtools and BCFtools. Gigascience 2021, 10, doi:10.1093/gigascience/giab008.

24. Weissensteiner, H.; Forer, L.; Fuchsberger, C.; Schöpf, B.; Kloss-Brandstätter, A.; Specht, G.; Kronenberg, F.; Schönherr, S. mtDNA-Server: next-generation sequencing data analysis of human mitochondrial DNA in the cloud. Nucleic Acids Res 2016, 44, W64–69, doi:10.1093/nar/gkw247.

25. Mbatchou, J.; Barnard, L.; Backman, J.; Marcketta, A.; Kosmicki, J.A.; Ziyatdinov, A.; Benner, C.; O’Dushlaine, C.; Barber, M.; Boutkov, B.; et al. Computationally efficient whole-genome regression for quantitative and binary traits. Nat Genet 2021, 53, 1097–1103, doi:10.1038/s41588-021-00870-7.

26. Manichaikul, A.; Mychaleckyj, J.C.; Rich, S.S.; Daly, K.; Sale, M.; Chen, W.M. Robust relationship inference in genome-wide association studies. Bioinformatics 2010, 26, 2867–2873, doi:10.1093/bioinformatics/btq559.

27. Gogarten, S.M.; Sofer, T.; Chen, H.; Yu, C.; Brody, J.A.; Thornton, T.A.; Rice, K.M.; Conomos, M.P. Genetic association testing using the GENESIS R/Bioconductor package. Bioinformatics 2019, 35, 5346–5348, doi:10.1093/bioinformatics/btz567.

28. Gogarten, S.M.; Zheng, X.; Stilp, A. SeqVarTools: tools for variant data. 2023, doi:10.18129/B9.bioc.SeqVarTools.

29. Wright, M.N.; Ziegler, A. ranger: a fast implementation of random forests for high dimensional data in C++ and R. J Stat Softw 2017, 77, doi:10.18637/jss.v077.i01.

30. Watson, D.S.; Wright, M.N. Testing conditional independence in supervised learning algorithms. Mach Learn 2021, 110, 2107–2129, doi:10.1007/s10994-021-06030-6.

31. Lang, M.; Binder, M.; Richter, J.; Schratz, P.; Pfisterer, F.; Coors, S.; Au, Q.; Casalicchio, G.; Kotthoff, L.; Bischl, B. mlr3: a modern object-oriented machine learning framework in R. J Open Source Softw 2019, 4, 1903, doi:10.21105/joss.01903.

32. R Core Team. R: A Language and Environment for Statistical Computing. Available online: https://www.R-project.org/ (accessed on 13.03.2024).

33. Landau, W.M. The targets R package: a dynamic make-like function-oriented pipeline toolkit for reproducibility and high-performance computing. J Open Source Softw 2019, 6, 2959, doi:10.21105/joss.02959.

34. Wickham, H. ggplot2: Elegant Graphics for Data Analysis. Springer-Verlag New York 2016.

35. Solomon, T.; Smith, E.N.; Matsui, H.; Braekkan, S.K.; Consortium, I.; Wilsgaard, T.; Njolstad, I.; Mathiesen, E.B.; Hansen, J.B.; Frazer, K.A. Associations between common and rare exonic genetic variants and serum levels of 20 cardiovascular-related proteins: the Tromsø Study. Circ Cardiovasc Genet 2016, 9, 375–383, doi:10.1161/CIRCGENETICS.115.001327.

36. Schachtl-Riess, J.F.; Kheirkhah, A.; Grüneis, R.; Di Maio, S.; Schoenherr, S.; Streiter, G.; Losso, J.L.; Paulweber, B.; Eckardt, K.U.; Köttgen, A.; et al. Frequent LPA KIV-2 variants lower lipoprotein(a) concentrations and protect against coronary artery disease. J Am Coll Cardiol 2021, 78, 437–449, doi:10.1016/j.jacc.2021.05.037.

37. Kraft, H.G.; Lingenhel, A.; Pang, R.W.; Delport, R.; Trommsdorff, M.; Vermaak, H.; Janus, E.D.; Utermann, G. Frequency distributions of apolipoprotein(a) kringle IV repeat alleles and their effects on lipoprotein(a) levels in Caucasian, Asian, and African populations: the distribution of null alleles is non-random. Eur J Hum Genet 1996, 4, 74–87, doi:10.1159/000472175.

38. van der Hoek, Y.Y.; Wittekoek, M.E.; Beisiegel, U.; Kastelein, J.J.; Koschinsky, M.L. The apolipoprotein(a) kringle IV repeats which differ from the major repeat kringle are present in variably-sized isoforms. Hum Mol Genet 1993, 2, 361–366, doi:10.1093/hmg/2.4.361.

