## Supplementary Figures for "Comprehensive analysis of the genetic variation in the *LPA* gene from short-read sequencing"

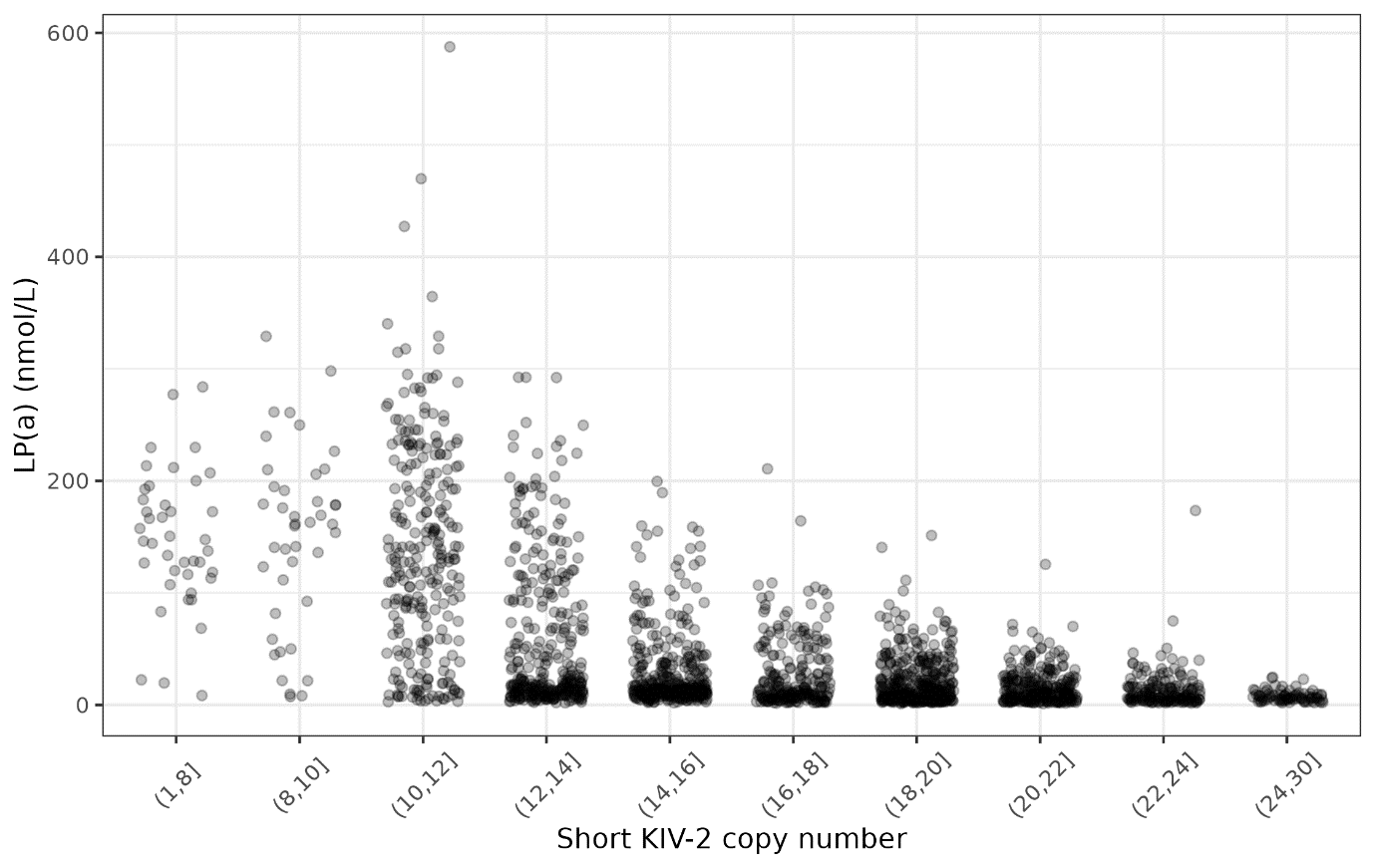

**Figure S1.** Scatterplot for LP(a) and number of KIV-2 repeats on the short allele (n = 4861). Number of KIV-2 repeats were binned so that there are at least 30 samples in each bin. Horizontal jittering was added to prevent overlapping between points.

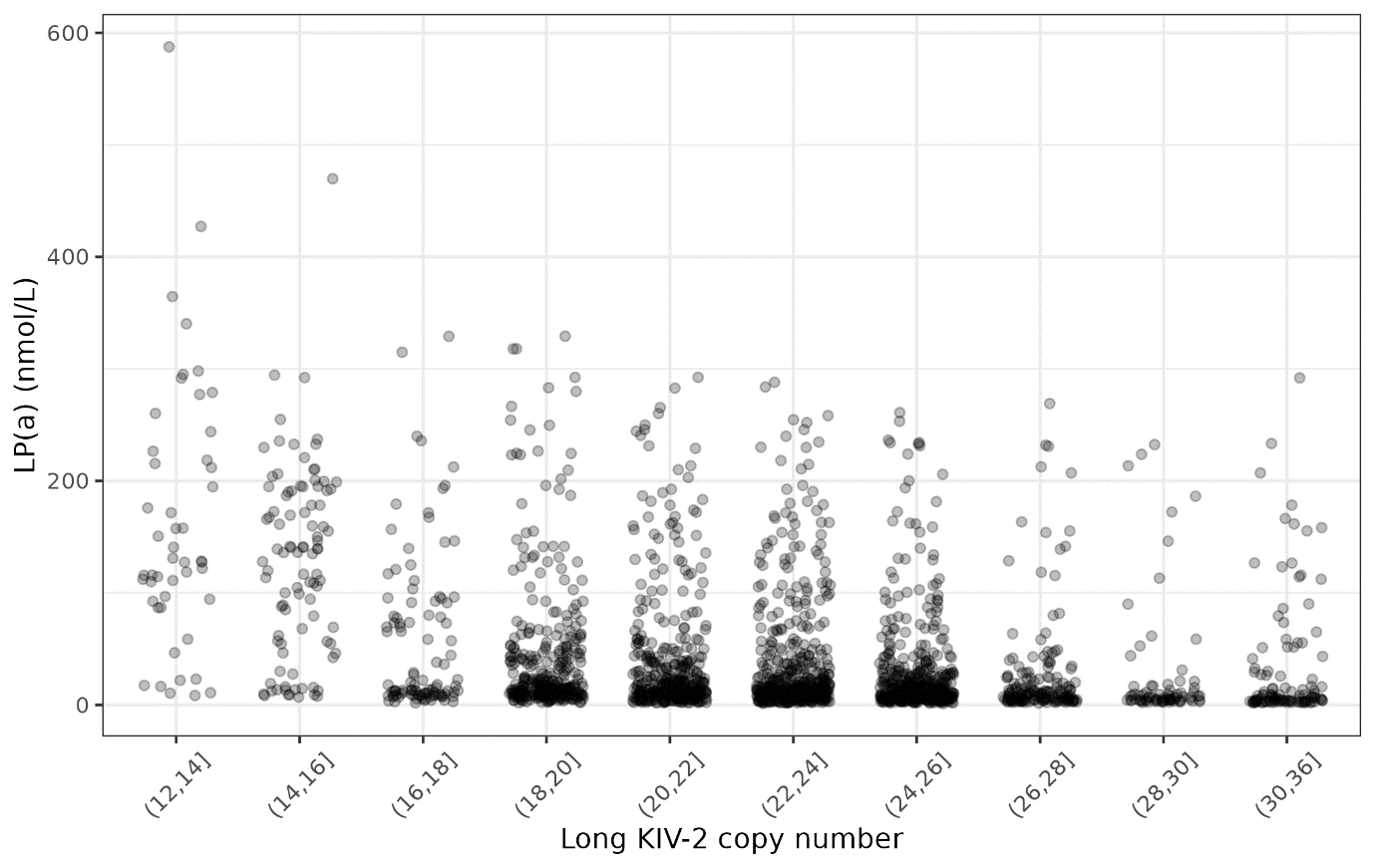

**Figure S2.** Scatterplot for LP(a) and number of KIV-2 repeats on the long allele (n = 4861). Number of KIV-2 repeats were binned so that there are at least 30 samples in each bin. Horizontal jittering was added to prevent overlapping between points.

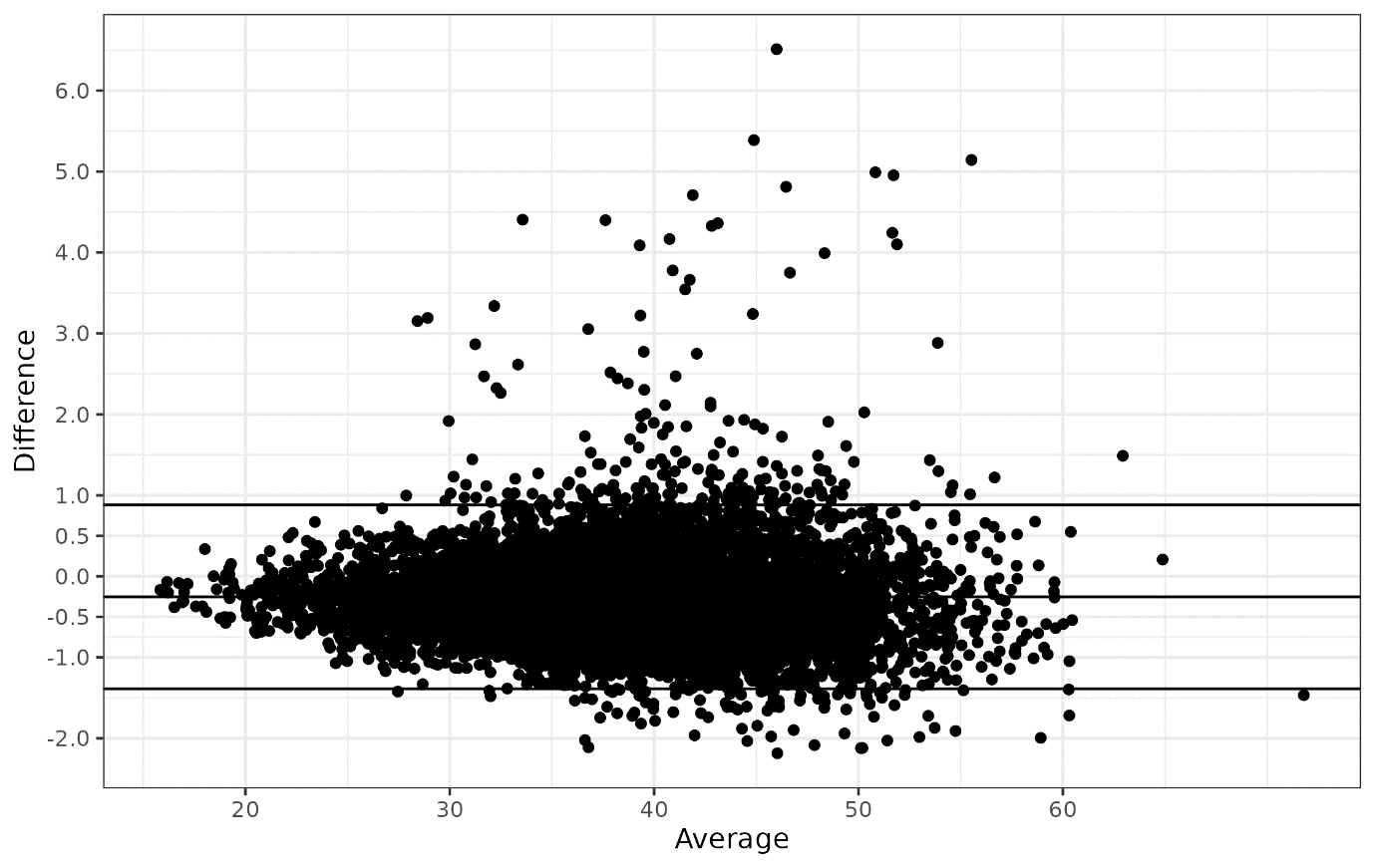
 **Figure S3.** Bland-Altman plot with mean and 95% confidence interval for total number of KIV-2 repeats estimated by the DRAGEN LPA Caller and read depth based CN estimator (CNE) (n = 8351).

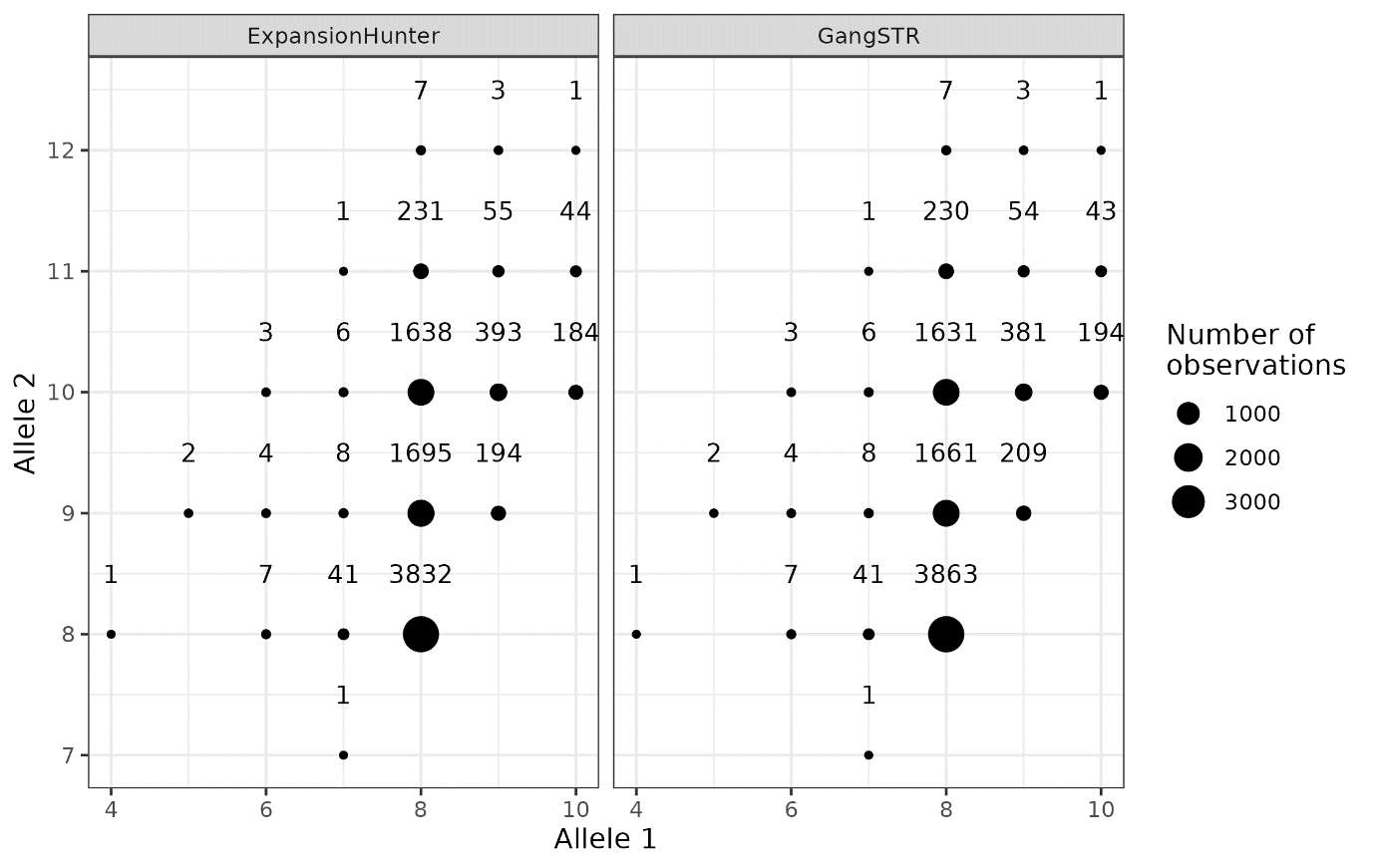

**Figure S4.** Dotplot per number of subjects per genotype estimated by ExpansionHunter and GangSTR.

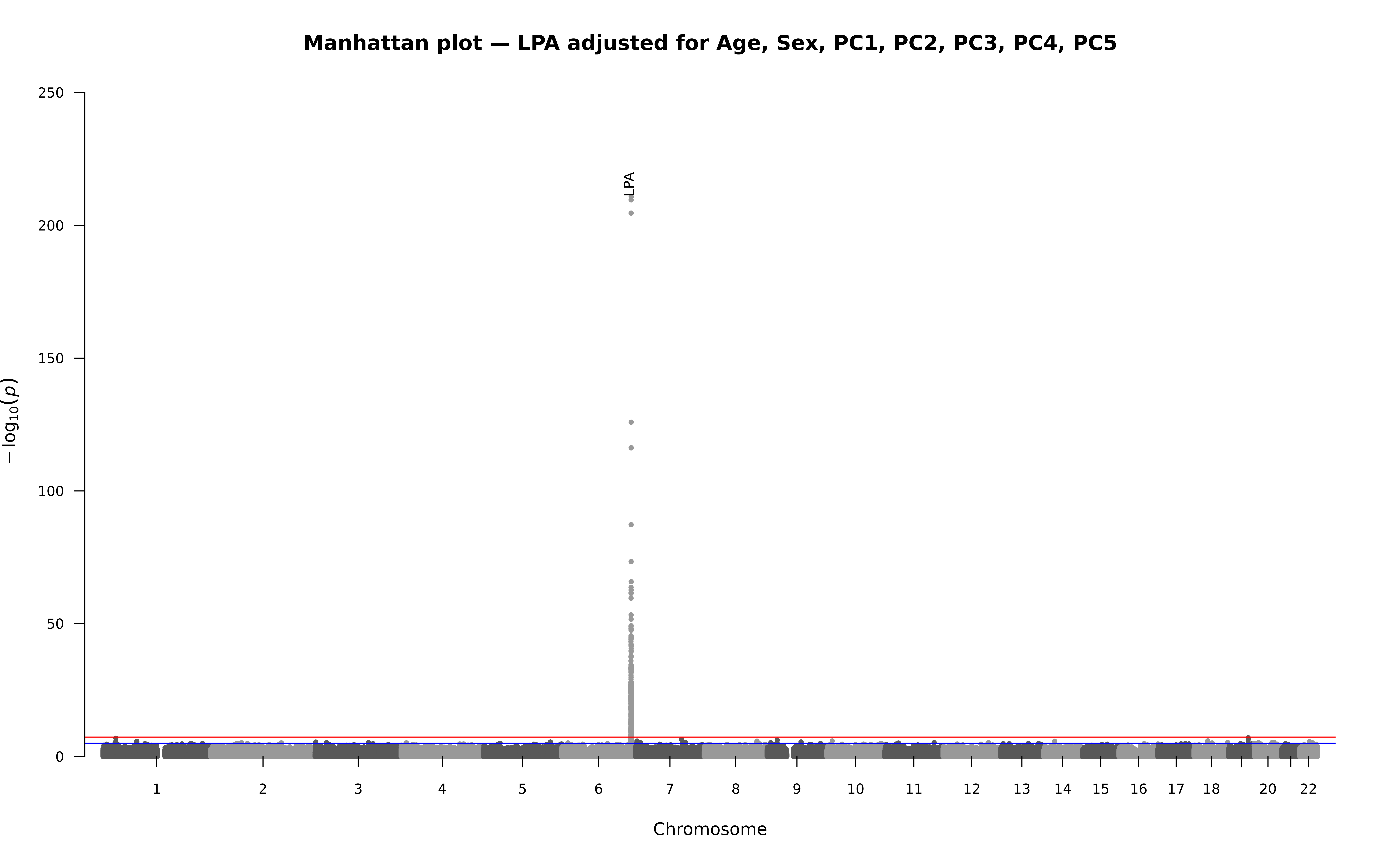

**Figure S5.** Manhattan plot from genome-wide association analysis (GWAS) for LP(a) concentration.

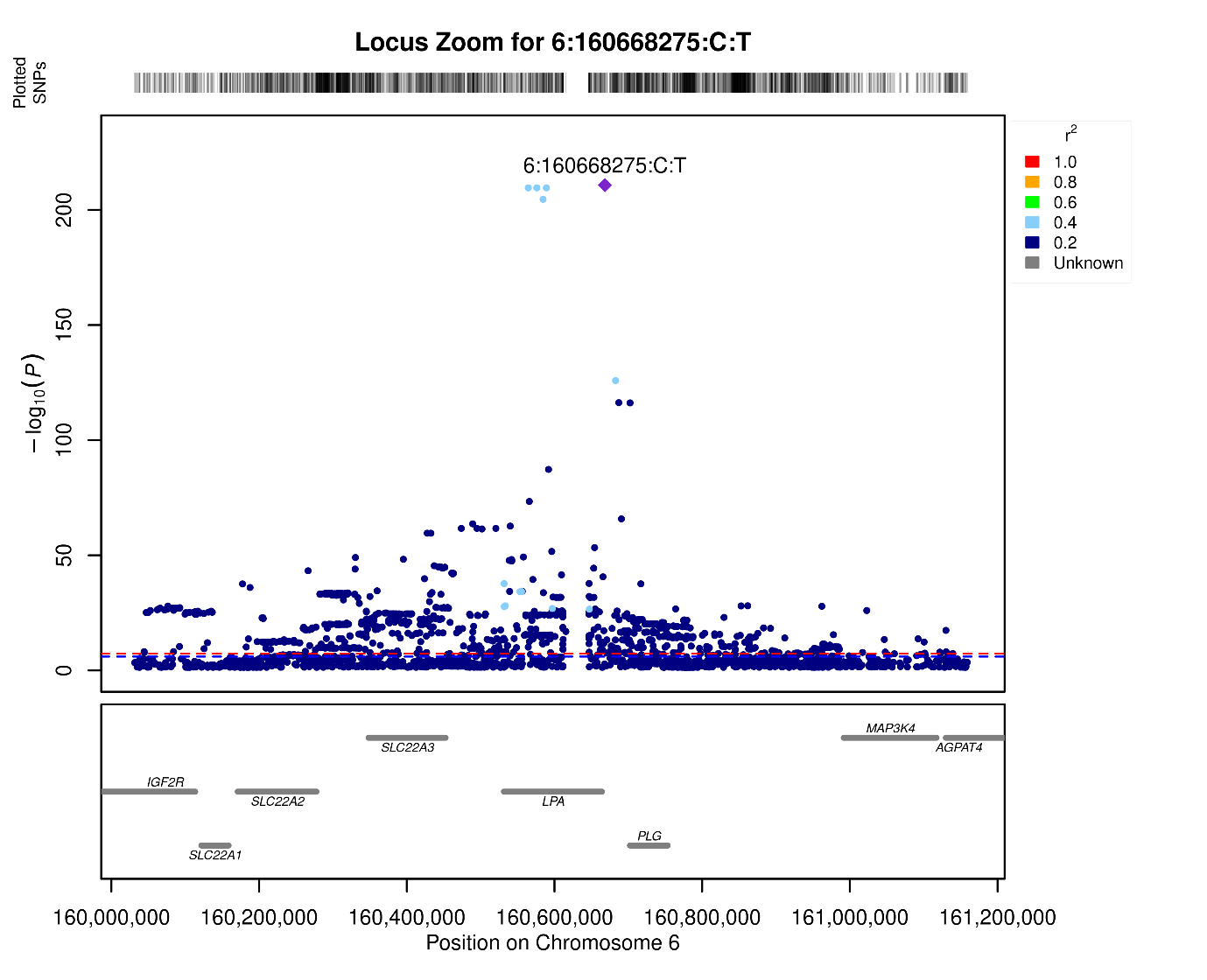
 **Figure S6.** Locus-zoom plot for the LPA gene with 500kb padding.
