## Supplementary R Code for "Comprehensive analysis of the genetic variation in the *LPA* gene from short-read sequencing": _targets.R

```
library(targets)
library(crew.cluster)
library(crew)

# Source file with packages
source("packages.R")

# Source functions
source("R/prepare_input.R")
source("R/cpi.R")
source("R/crossvalidate_performance.R")
source("R/crossvalidate_performance_short.R")
source("R/crossvalidate_performance_herit.R")

# Source plan
source("plans/input_file_list.R")
source("plans/data_preparation.R")
source("plans/model_dragen_total.R")
source("plans/model_dragen_shortlong.R")
source("plans/performance_models.R")

# Setup jobs which run locally
controller_local <- crew_controller_local(
  name = "my_local_controller",
  workers = 3,
  seconds_idle = 10
)

# Setup SLURM template file, maximum 3 SLURM jobs at once
controller_slurm <- crew_controller_slurm(
  name = "my_SLURM_controller",
  workers = 3,
  script_lines = c("source /software/modules/init/bash",
                   "ml R/4.3.2"), 
  seconds_interval = 5,
  slurm_cpus_per_task = 120,
  slurm_memory_gigabytes_per_cpu = 4,
  slurm_partition = "nodes",
  seconds_idle = 10
)

# Set default storage format to qs and specify controller groups
tar_option_set(
  format = "qs",
  controller = crew_controller_group(controller_local, controller_slurm),
  resources = tar_resources(
    crew = tar_resources_crew(controller = "my_local_controller")
  )
)

# 
list(`define_files`,
     `data_preparation`,
     `model_DRAGEN_shortlong`,
     `model_DRAGEN_total`,
     `full_model_metrics`,
     `medium_model_metrics`,
     `small_model_metrics`,
     `full_model_metrics_short`,
     `medium_model_metrics_short`,
     `small_model_metrics_short`,
     `full_model_metrics_herit`,
     `medium_model_metrics_herit`,
     `small_model_metrics_herit`
)
```
