## Supplementary R Code for "Comprehensive analysis of the genetic variation in the *LPA* gene from short-read sequencing": packages.R

```
tar_option_set(
  packages = c(
    "tidyverse", 
    "SeqArray", 
    "SeqVarTools", 
    "cpi",
    "caret",
    "foreach",
    "ranger",
    "mlr3", 
    "mlr3learners", 
    "mlr3tuning",
    "doParallel")
  )
```
