## Supplementary R Code for "Comprehensive analysis of the genetic variation in the *LPA* gene from short-read sequencing": data_preparation.R

```
"data_preparation" = list(
  # Load phenotypes file
  tar_target(phenotypes,
             get_phenotypes(file_phenotypes)),
  # Regenie GWAS results from Roche assay
  tar_target(GWAS, 
             get_roche_gwas_snvs(file_regenie_roche, file_GDS)),
  # Get SNVs within KIV-2 repeat
  tar_target(KIV2_SNVs, 
             get_snvs_in_KIV2(file_sample_names_kiv2_snvs, file_kiv2_snvs)),
  # Get number of PNRs from GangSTR
  tar_target(PNR_GangSTR, 
             get_number_pnr_gangstr(file_sample_names, file_gangstr)),
  # Get number of PNRs from ExpansionHunter
  tar_target(PNR_ExpHunt, 
             get_number_pnr_exphunt(file_sample_names, file_exphunt)),
  # Get KIV-2 copy numbers from DRAGEN
  tar_target(KIV2_CopyNumbers_DRAGEN,
             get_kiv2_copy_number()),
  # Get KIV-2 copy numbers from CNV Estimator
  tar_target(KIV2_CopyNumbers_CNV_Estimator,
             get_kiv2_copy_number_cnv_estimator()),
  # Merge everything into dataframe
  tar_target(complete_dataframe,
             merge_all(phenotypes, 
                       KIV2_CopyNumbers_DRAGEN,
                       GWAS, 
                       KIV2_SNVs,
                       PNR_GangSTR)),
  # Create STR dataframe for GangSTR / ExpansionHunter comparison
  tar_target(str_df,
             create_str_df(PNR_GangSTR, PNR_ExpHunt))
)
```
