## Supplementary R Code for "Comprehensive analysis of the genetic variation in the *LPA* gene from short-read sequencing": input_file_list.R

```
# This file defines all the input files used for the analyses

"define_files" = list(
  tar_target(file_regenie_roche,
             <REGENIE_FILE>,
             format = "file"),
  tar_target(file_GDS,
             <GDS_FILE_GENOTYPE>,
             format = "file"),
  tar_target(file_sample_names_kiv2_snvs,
             <SAMPLE_NAMES_FILE>,
             format = "file"),
  tar_target(file_kiv2_snvs,
             <KIV2_CALLER_FILE>,
             format = "file"),
  tar_target(file_sample_names,
             <STR_SAMPLE_NAMES_FILE>,
             format = "file"),
  tar_target(file_gangstr,
             <GANGSTR_FILE>,
             format = "file"),
  tar_target(file_exphunt,
             <EXPANSIONHUNTER_FILE>,
             format = "file"),
  tar_target(file_phenotypes,
             <PHENOTYPES_FILE>,
             format = "file")
)
```
