## Supplementary R Code for "Comprehensive analysis of the genetic variation in the *LPA* gene from short-read sequencing": model_dragen_shortlong.R

```
# Model with DRAGEN LPA caller haplotype-resolved KIV-2 copy number
# KIV2_SHORT and KIV2_LONG

"model_DRAGEN_shortlong" = list(
  tar_target(model_dragen_shortlong,
             create_dragen_shortlong_KIV2_dataframe(complete_dataframe)),

  # Get best hyperparameters for ranger
  tar_target(optimal_ranger_hyperparameters_model_dragen_shortlong,
             get_best_hyperparameter_ranger(model_dragen_shortlong),
             resources = tar_resources(
               crew = tar_resources_crew(controller = "my_SLURM_controller")
             )
  ),
  tar_target(cpi_ranger_dragen_shortlong,
             calculate_cpi_ranger(model_dragen_shortlong),
             resources = tar_resources(
               crew = tar_resources_crew(controller = "my_SLURM_controller")
             )
  )
)
```
