## Supplementary R Code for "Comprehensive analysis of the genetic variation in the *LPA* gene from short-read sequencing": model_dragen_total.R

```
# Model with DRAGEN LPA caller only total KIV-2 CN

"model_DRAGEN_total" = list(
  tar_target(model_dragen_total,
             create_dragen_total_KIV2_dataframe(complete_dataframe)),
  # Get best hyperparameters for ranger
  tar_target(optimal_ranger_hyperparameters_model_dragen_total,
             get_best_hyperparameter_ranger(model_dragen_total),
             resources = tar_resources(
               crew = tar_resources_crew(controller = "my_SLURM_controller")
             )
  ),
  #Run CPI for ranger
  tar_target(cpi_ranger_dragen_total,
             calculate_cpi_ranger(model_dragen_total),
             resources = tar_resources(
               crew = tar_resources_crew(controller = "my_SLURM_controller")
             )
  )
)
```
