## Supplementary R Code for "Comprehensive analysis of the genetic variation in the *LPA* gene from short-read sequencing": performance_models.R

```
# Evaluate performance between total and short + long number of KIV-2 repeats
"full_model_metrics" = list(
  tar_target(metrics_full_model,
             big_model_performance(model_dragen_shortlong, model_dragen_total),
             resources = tar_resources(
               crew = tar_resources_crew(controller = "my_SLURM_controller")
             ))
)

"medium_model_metrics" = list(
  tar_target(metrics_medium_model,
             medium_model_performance(model_dragen_shortlong, model_dragen_total),
             resources = tar_resources(
               crew = tar_resources_crew(controller = "my_SLURM_controller")
             ))
)

"small_model_metrics" = list(
  tar_target(metrics_small_model,
             small_model_performance(model_dragen_shortlong, model_dragen_total),
             resources = tar_resources(
               crew = tar_resources_crew(controller = "my_SLURM_controller")
             ))
)

# Evaluate performance between short and short + long number of KIV-2 repeats
"full_model_metrics_short" = list(
  tar_target(metrics_full_model_short,
             big_model_performance_short(model_dragen_shortlong, model_dragen_total),
             resources = tar_resources(
               crew = tar_resources_crew(controller = "my_SLURM_controller")
             ))
)

"medium_model_metrics_short" = list(
  tar_target(metrics_medium_model_short,
             medium_model_performance_short(model_dragen_shortlong, model_dragen_total),
             resources = tar_resources(
               crew = tar_resources_crew(controller = "my_SLURM_controller")
             ))
)

"small_model_metrics_short" = list(
  tar_target(metrics_small_model_short,
             small_model_performance_short(model_dragen_shortlong, model_dragen_total),
             resources = tar_resources(
               crew = tar_resources_crew(controller = "my_SLURM_controller")
             ))
)

"full_model_metrics_herit" = list(
  tar_target(metrics_full_model_herit,
             big_model_performance_sex_age(model_dragen_shortlong, model_dragen_total),
             resources = tar_resources(
               crew = tar_resources_crew(controller = "my_SLURM_controller")
             ))
)

"medium_model_metrics_herit" = list(
  tar_target(metrics_medium_model_herit,
             medium_model_performance_sex_age(model_dragen_shortlong, model_dragen_total),
             resources = tar_resources(
               crew = tar_resources_crew(controller = "my_SLURM_controller")
             ))
)

"small_model_metrics_herit" = list(
  tar_target(metrics_small_model_herit,
             small_model_performance_sex_age(model_dragen_shortlong, model_dragen_total),
             resources = tar_resources(
               crew = tar_resources_crew(controller = "my_SLURM_controller")
             ))
)
```
