## Supplementary R Code for "Comprehensive analysis of the genetic variation in the *LPA* gene from short-read sequencing": cpi.R

```
################################################################################
######################### CPI calculation for ranger ###########################
################################################################################

calculate_cpi_ranger <- function(dataframe_model){
  # Fix multithread seed
  set.seed(42, "L'Ecuyer-CMRG")
  
  # Allocate 120 cores for CPI
  doParallel::registerDoParallel(120)
  
  # Remove variables with zero variance
  zero_var_cols <- nearZeroVar(x = dataframe_model, uniqueCut = 0)
  dataframe_model <- dataframe_model[,-zero_var_cols]
  
  # Run CPI with ranger, always select Sex and Age
  ranger_cpi <- cpi(
    task = as_task_regr(LP_A_Roche_log ~ ., 
                        data = dataframe_model),
    learner = lrn("regr.ranger", 
                  mtry.ratio = 0.725,
                  max.depth = 10,
                  min.node.size = 5,
                  always.split.variables = c("Sex", "Age")),
    resampling = rsmp("cv", folds = 10),
    verbose = TRUE
  )
  
  ranger_cpi <- as_tibble(ranger_cpi) %>%
    mutate(abs_change = abs(estimate)) %>%
    arrange(desc(abs_change))
  
  return (ranger_cpi)
  
}

################################################################################
####################### HPO for ranger #########################################
################################################################################

get_best_hyperparameter_ranger <- function(dataframe_model){
  # Fix multithread seed
  set.seed(42, "L'Ecuyer-CMRG")
  
  future::plan(list(
    future::tweak("multisession", workers = 10),
    future::tweak("multisession", workers = 12)
  ))
  
  # Create regression task
  task <- as_task_regr(LP_A_Roche_log ~ ., 
                       data = dataframe_model)
  
  # Create ranger regression learner
  learner <- lrn("regr.ranger",
                 max.depth = to_tune(3, 10),
                 num.trees = 1000,
                 mtry.ratio = to_tune(0.05, 0.95),
                 min.node.size = to_tune(5, 200),
                 always.split.variables = c("Sex", "Age")
                 
  )
  
  # Grid search tuner, resolution specifies how many combinations
  tnr_gridsearch = tnr("grid_search", 
                       resolution = 5, 
                       batch_size = 10)
  
  generate_design_grid(learner$param_set$search_space(), 
                       resolution = 5)
  
  # 10 fold CV for both inner and outer loop
  rsmp_cv10 <- rsmp("cv", 
                    folds = 10)
  
  # No stop criteria, explore all combinations
  trm = trm("none")
  
  # Use MSE measure as objective function to minimize
  mse = msr("regr.mse")
  
  # Autotune
  at = auto_tuner(tuner = tnr_gridsearch,
                  learner = learner,
                  resampling = rsmp_cv10,
                  measure = mse,
                  terminator = trm
  )
  
  # In the inner loop, find the best hyperparameters and
  # in the outer loop, test the model
  rr = resample(task = task,
                learner = at,
                resampling = rsmp_cv10,
                store_models = TRUE)
  
  return (rr)
}
```
