## Supplementary R Code for "Comprehensive analysis of the genetic variation in the *LPA* gene from short-read sequencing": crossvalidate_performance.R

```
library(mlr3)
library(mlr3learners)
library(mlr3tuning)
library(targets)
library(tidyverse)

# Small model with only variables from allele-specific KIV2-CN CPI
small_model_performance <- function(shortlong_dataframe, total_dataframe){
  set.seed(42, "L'Ecuyer-CMRG")
  
  lgr::get_logger("mlr3")$set_threshold("debug")
  
  future::plan(list(
    future::tweak("multisession", workers = 20),
    future::tweak("multisession", workers = 6)
  ))
  
  model_shortlong <- shortlong_dataframe
  model_total <- total_dataframe
  
  ################################################################################
  ############################# Specify tasks ####################################
  ################################################################################
  
  task_shortlong = as_task_regr(LP_A_Roche_log ~ KIV2_SHORT + KIV2_LONG + 
                                  SNP_4925_G_A + SNV_160585045 + 
                                  Age + Sex,
                                data = model_shortlong,
                                id = "Shortlong")
  
  task_total = as_task_regr(LP_A_Roche_log ~ KIV2_TOTAL + 
                              SNP_4925_G_A + SNV_160585045 + 
                              Age + Sex,
                            data = model_total,
                            id = "Total")
  
  tasks = as_tasks(c(task_shortlong, task_total))
  
  ################################################################################
  ######################## Specify resampling ####################################
  ################################################################################
  
  # 10-fold CV for both inner fold (HPO) and outer fold (measure performance)
  resampling = rsmp("cv", folds = 10)
  
  ################################################################################
  ########################## Specify learners ####################################
  ################################################################################

  learner_ranger = lrn("regr.ranger",
                       max.depth = to_tune(3, 10),
                       num.trees = to_tune(100, 1000),
                       mtry.ratio = 1,
                       min.node.size = to_tune(5, 20))
  
  # Grid search tuner, resolution specifies how many combinations
  tnr_gridsearch = tnr("grid_search", 
                       resolution = 5, 
                       batch_size = 10)
  
  # generate_design_grid(learner$param_set$search_space(), 
  #                      resolution = 5)
  
  # No stop criteria, explore all combinations
  trm = trm("none")
  
  # Use MSE measure as objective function to minimize
  mse = msr("regr.mse")
  
  # Autotune ranger
  at = auto_tuner(tuner = tnr_gridsearch,
                  learner = learner_ranger,
                  resampling = resampling,
                  measure = mse,
                  terminator = trm
  )

  design = benchmark_grid(tasks,
                          at, 
                          resampling)
  
  # Set store_models to TRUE if you want to get a look at the fitted models
  bmr = benchmark(design, store_models = TRUE)
  
  return (bmr)  
}

# Medium  model with variables from total KIV2-CN CPI
medium_model_performance <- function(shortlong_dataframe, total_dataframe){
  set.seed(42, "L'Ecuyer-CMRG")
  
  lgr::get_logger("mlr3")$set_threshold("debug")
  
  future::plan(list(
    future::tweak("multisession", workers = 20),
    future::tweak("multisession", workers = 6)
  ))
  
  model_shortlong <- shortlong_dataframe
  model_total <- total_dataframe
  
  ################################################################################
  ############################# Specify tasks ####################################
  ################################################################################
  
  task_shortlong = as_task_regr(LP_A_Roche_log ~ KIV2_SHORT + KIV2_LONG +
                                  SNP_4925_G_A + SNV_160585045 + SNV_160868900 +
                                  SNV_160898903 + SNV_160487052 + Age + Sex,
                                data = model_shortlong,
                                id = "Shortlong")

  task_total = as_task_regr(LP_A_Roche_log ~ KIV2_TOTAL + 
                              SNP_4925_G_A + SNV_160585045 + SNV_160868900 +
                              SNV_160898903 + SNV_160487052 + Age + Sex,
                            data = model_total,
                            id = "Total")
  
  tasks = as_tasks(c(task_shortlong, task_total))
  
  ################################################################################
  ######################## Specify resampling ####################################
  ################################################################################
  
  # 10-fold CV for both inner fold (HPO) and outer fold (measure performance)
  resampling = rsmp("cv", folds = 10)
  
  ################################################################################
  ########################## Specify learners ####################################
  ################################################################################
  
  learner = lrn("regr.ranger",
                max.depth = to_tune(3, 10),
                num.trees = to_tune(100, 1000),
                mtry.ratio = 1,
                min.node.size = to_tune(5, 20))
  
  # Grid search tuner, resolution specifies how many combinations
  tnr_gridsearch = tnr("grid_search", 
                       resolution = 5, 
                       batch_size = 10)
  
  generate_design_grid(learner$param_set$search_space(), 
                       resolution = 5)
  
  # No stop criteria, explore all combinations
  trm = trm("none")
  
  # Use MSE measure as objective function to minimize
  mse = msr("regr.mse")
  
  # Autotune
  at = auto_tuner(tuner = tnr_gridsearch,
                  learner = learner,
                  resampling = resampling,
                  measure = mse,
                  terminator = trm
  )
  
  design = benchmark_grid(tasks, at, resampling)
  
  # Set store_models to TRUE if you want to get a look at the fitted models
  bmr = benchmark(design, store_models = FALSE)
  
  return (bmr)  
}


big_model_performance <- function(shortlong_dataframe, total_dataframe){
  set.seed(42, "L'Ecuyer-CMRG")
  
  lgr::get_logger("mlr3")$set_threshold("debug")
  
  future::plan(list(
    future::tweak("multisession", workers = 20),
    future::tweak("multisession", workers = 6)
  ))
  
  model_shortlong <- shortlong_dataframe
  model_total <- total_dataframe
  
  ##############################################################################
  ############################# Specify tasks ##################################
  ##############################################################################
  
  task_shortlong = as_task_regr(LP_A_Roche_log ~ .,
                                data = model_shortlong,
                                id = "Shortlong")
  
  task_total = as_task_regr(LP_A_Roche_log ~ .,
                            data = model_total,
                            id = "Total")
  
  tasks = as_tasks(c(task_shortlong, task_total))
  
  ##############################################################################
  ######################## Specify resampling ##################################
  ##############################################################################
  
  # 10-fold CV for both inner fold (HPO) and outer fold (measure performance)
  resampling = rsmp("cv", folds = 10)
  
  ##############################################################################
  ########################## Specify learners ##################################
  ##############################################################################
  
  # Create ranger regression learner
  learner <- lrn("regr.ranger",
                 max.depth = to_tune(3, 10),
                 num.trees = 1000,
                 mtry.ratio = to_tune(0.05, 0.95),
                 min.node.size = to_tune(5, 200),
                 always.split.variables = c("Sex", "Age"))
  
  # Grid search tuner, resolution specifies how many combinations
  tnr_gridsearch = tnr("grid_search", 
                       resolution = 5, 
                       batch_size = 10)
  
  generate_design_grid(learner$param_set$search_space(), 
                       resolution = 5)
  
  # No stop criteria, explore all combinations
  trm = trm("none")
  
  # Use MSE measure as objective function to minimize
  mse = msr("regr.mse")
  
  # Autotune
  at = auto_tuner(tuner = tnr_gridsearch,
                  learner = learner,
                  resampling = resampling,
                  measure = mse,
                  terminator = trm
  )
  
  design = benchmark_grid(tasks, at, resampling)
  
  # Set store_models to TRUE if you want to get a look at the fitted models
  bmr = benchmark(design, store_models = FALSE)
  
  return (bmr)  

}
```
