## Supplementary R Code for "Comprehensive analysis of the genetic variation in the *LPA* gene from short-read sequencing": prepare_input.R

```
# This file defines all the functions used to create the dataframe

# Parse GWAS information from REGENIE
get_roche_gwas_snvs <- function(file_regenie_roche, file_GDS) {
  # Get genomic position of LPA gene
  ensembl <- biomaRt::useMart("ensembl")
  ensembl <- biomaRt::useDataset("hsapiens_gene_ensembl", mart = ensembl)
  genomic_pos <- biomaRt::getBM(attributes = c("start_position", "end_position"),
                                filters = "hgnc_symbol",
                                values= "LPA",
                                mart = ensembl)
  # Up- and downstream range
  SNP_range <- 500000
  significant_snvs <- read_delim(file_regenie_roche, delim = " ") %>% 
    filter(LOG10P >= -log10(5e-8),
           (str_length(ALLELE0) + str_length(ALLELE1)) == 2,
           N == 4803,
           between(GENPOS,
                   as.numeric(genomic_pos["start_position"] - SNP_range),
                   as.numeric(genomic_pos["end_position"]+ SNP_range))) %>% 
    pull(ID)
  
  # Load the GDS file
  genofile <- seqOpen(file_GDS)
  
  # Only keep SNVs from GWAS
  seqSetFilterAnnotID(genofile, significant_snvs)
  
  # Get the dosage for the alt allele
  alt_dosage <- altDosage(genofile)
  
  annotations <- paste0("SNV_", seqGetData(genofile, "position"))
  
  df <- as_tibble(alt_dosage, rownames = "sample_name_vc")
  colnames(df) <- c("sample_name_vc", annotations)
  
  # Set missing genotypes to 0/0
  df <- df %>% 
    replace(is.na(.), 0)
  
  return (df)
}

# Parse information from specialized KIV-2 SNV caller
get_snvs_in_KIV2 <- function(file_sample_names_kiv2_snvs, file_kiv2_snvs){
  sample_names <- read_tsv(file_sample_names_kiv2_snvs,
                           col_names = "sample_name_vc") %>% 
    pull(sample_name_vc)
  
  KIV2_snps <- read_tsv(file_kiv2_snvs,
                        col_names = c("CHROM", "POS", "REF", "ALT", sample_names)) %>%
    pivot_longer(cols = starts_with("HH2020_"), 
                 names_to = "sample_name_vc",
                 values_to = "Genotype") %>% 
    mutate(SNP_carriers = if_else(Genotype == "0/0", 0, 1)) %>%
    select(-c(Genotype, CHROM)) %>% 
    pivot_wider(names_from = POS:ALT,
                values_from = SNP_carriers, 
                names_prefix = "SNP_")
  
  
  return (KIV2_snps)
}

get_number_pnr_gangstr <- function(file_sample_names, file_gangstr){
  samples <- read_lines(file_sample_names)
  
  gangstr <- read_tsv(file_gangstr,
                      col_names = c("CHROM", "POS", "RU", samples, "tmp"),
                      col_types = paste(rep("c", (length(samples) + 3)), collapse = "")) %>%
    filter(RU == "aataa", POS == "160665585") %>% 
    pivot_longer(cols = starts_with("HH2020_"), 
                 names_to = "sample_name_vc", values_to = "REPCN") %>% 
    select(-tmp) %>% 
    separate(col = REPCN, into = c("A1", "A2"), sep = ",", convert = TRUE) %>%
    rowwise() %>% 
    mutate(A1_min = as.numeric(min(A1, A2)),
           A2_max = as.numeric(max(A1, A2))) %>% 
    select(-c(A1, A2, CHROM, POS, RU)) %>% 
    rename(PNR_A1 = A1_min,
           PNR_A2 = A2_max)
  
  return (gangstr)
}

get_number_pnr_exphunt <-  function(file_sample_names, file_exphunt){
  samples <- read_lines(file_sample_names)
  
  exphunt <- read_tsv(file_exphunt,
                      col_names = c("CHROM", "POS", "RU", samples, "tmp"),
                      col_types = paste(rep("c", (length(samples) + 3)), collapse = "")) %>% 
    filter(RU == "AATAA", POS == "160665584") %>% 
    pivot_longer(cols = starts_with("HH2020_"),
                 names_to = "sample_name_vc", values_to = "REPCN") %>%
    select(-tmp) %>% 
    separate(col = REPCN, into = c("A1", "A2"), sep = "/", convert = TRUE) %>%
    rowwise() %>%
    mutate(A1_min = as.numeric(min(A1, A2)),
           A2_max = as.numeric(max(A1, A2))) %>%
    select(-c(A1, A2, CHROM, POS, RU)) %>%
    rename(PNR_A1 = A1_min,
           PNR_A2 = A2_max)
  
  return (exphunt)
  
}

# Read the file with phenotypes (LP(a), Age and Sex) and the file with
# the PC coefficients
get_phenotypes <- function(file_phenotype, file_pca){
  # Remove two samples with LP(a) of 0
  pheno <- read_csv(file_phenotype) %>% 
    select(sample_name_vc, include_msVCF, LP_A_Roche, HCH_SVAGE0001, HCH_SVSEX0001) %>% 
    filter(include_msVCF == TRUE,
           LP_A_Roche != 0) %>% 
    rename(Age = HCH_SVAGE0001,
           Sex = HCH_SVSEX0001) %>% 
    select(-c(include_msVCF))
  
  return (pheno)
}

## Get the KIV-2 CN from the DRAGEN json file for all 8351 samples and
## add KIV2_SHORT and KIV2_LONG variable
get_kiv2_copy_number <- function(){
  create_tibble_from_json <- function(file_path){
    tmp <- jsonlite::fromJSON(file_path)
    df <- tibble(sample_name_vc = tmp$Sample,
                 KIV2_TOTAL = tmp$`Target Calls`$`Kringle-IV-2 Copy Number`,
                 KIV2_REF = tmp$`Target Calls`$`REF Marker Allele Copy Number`,
                 KIV2_ALT = tmp$`Target Calls`$`ALT Marker Allele Copy Number`)
    
    return (df)
  }
  
  dirs <- Sys.glob(c("/cluster/storage/data/WGS_HCHS/phase1/call/DRAGEN_42_LPA_only/",
                     "/cluster/storage/data/WGS_HCHS/phase2/*/call/DRAGEN_42_LPA_only/"))
  
  json_files <- list.files(path = dirs,
                           pattern = "hg38_alt_aware-38c-adap-LPA_only.targeted.json",
                           recursive = TRUE,
                           full.names = TRUE)
  
  kiv2_copy_numbers_dragen <- lapply(json_files, create_tibble_from_json) %>% 
    bind_rows() %>% 
    rowwise() %>% 
    mutate(KIV2_SHORT = min(KIV2_REF, KIV2_ALT),
           KIV2_LONG = max(KIV2_REF, KIV2_ALT))
  
  return (kiv2_copy_numbers_dragen)
  
}

## Get the KIV-2 CN from the CNV Estimator normalized file
get_kiv2_copy_number_cnv_estimator <- function(){
  files <- list.files("/cluster/storage/scratch/rbetschart_tmp/CNV_estimator", 
                      pattern = "norm.txt.gz", 
                      full.names = TRUE)
  
  kiv2_copy_numbers_cnvestimator <- lapply(files, read_tsv, show_col_type = FALSE) %>% 
    bind_rows() %>% 
    mutate(sample_name_vc = sampleID,
           KIV2_TOTAL_CNV_Estimator = 6 * CN) %>% 
    select(sample_name_vc, KIV2_TOTAL_CNV_Estimator)
  
  return (kiv2_copy_numbers_cnvestimator)
}

merge_all <- function(dataframe_phenotypes, 
                      kiv2_copy_numbers_dragen,
                      dataframe_gwas, 
                      dataframe_kiv2_snvs, 
                      dataframe_gangstr){
  df <- purrr::reduce(list(dataframe_phenotypes,
                           kiv2_copy_numbers_dragen,
                           dataframe_gwas, 
                           dataframe_kiv2_snvs, 
                           dataframe_gangstr),
                      left_join) %>% 
    select(-contains(c("*", ","))) %>% 
    mutate(LP_A_Roche_log = log(LP_A_Roche))
  
  return (df)
}

create_str_df <- function(str_gangstr, str_exphunt){
  gangstr <- str_gangstr %>% 
    mutate(Algorithm = "GangSTR")
  
  exphunt <- str_exphunt %>% 
    mutate(Algorithm = "ExpHunt")
  
  df <- bind_rows(gangstr, exphunt)
  
  return (df)
  
}

# Create a dataframe with DRAGEN LPA caller KIV-2 copy numbers 
# KIV2_SHORT and KIV2_LONG
# Only keep haplotype resolved samples

create_dragen_shortlong_KIV2_dataframe <- function(dataframe_model){
  df <- dataframe_model %>% 
    filter(!is.na(KIV2_SHORT),
           !is.na(KIV2_LONG)) %>% 
    select(LP_A_Roche_log,
           starts_with("SNV_"),
           starts_with("SNP_"),
           starts_with("PNR_"),
           KIV2_SHORT, KIV2_LONG,
           Sex, Age)
  
  return (df)
}


# Create a dataframe with DRAGEN LPA caller KIV-2 copy numbers 
# KIV-2 TOTAL only
# Only keep haplotype resolved samples

create_dragen_total_KIV2_dataframe <- function(dataframe_model){
  df <- dataframe_model %>% 
    filter(!is.na(KIV2_REF),
           !is.na(KIV2_ALT)) %>% 
    select(LP_A_Roche_log,
           starts_with("SNV_"),
           starts_with("SNP_"),
           starts_with("PNR_"),
           KIV2_TOTAL,
           Sex, Age)
  
  return (df)
}
```
