## Supplementary R Code for "Comprehensive analysis of the genetic variation in the *LPA* gene from short-read sequencing": README.md

# Analysis for manuscript *Comprehensive analysis of the genetic variation in the* LPA *gene from short-read sequencing*

This folder contains R scripts used for the analysis, based on `targets`

To run the analysis, run `tar_make()`.

-   `packages.R` contains the list of packages used in the analyses

-   `targets.R` is the main file used by `targets`

-   R functions are stored under `R/`

    -   `prepare_input.R` prepares the input from GWAS, phenotypes, etc.

    -   `cpi.R` performs conditional predictive impact (CPI) based on optimized hyperparameters

    -   `crossvalidate_performance*.R` calculates the predictive performance for the three models

-   Target plans are stored under `plans/`

    -   `input_file_list.R` holds the file path for necessary files (GDS file with genotypes, file with phenotypes, etc.)

    -   `data_preparation.R` creates dataframes

    -   `model_dragen_*.R` holds the plans for total and allele-specific KIV-2 number models

    -   `performance_models.R` holds the plans to calculate the predictive performance across models
